# Extensive Healthy Donor Age/Gender Adjustments and Propensity Score Matching Reveal Physiology of Multiple Sclerosis through Immunophenotyping

**DOI:** 10.1101/2020.05.26.20113902

**Authors:** Paavali A. Hannikainen, Peter Kosa, Christopher Barbour, Bibiana Bielekova

**Author notes:** **Correspondence:** Bibiana Bielekova.

## Abstract

Quantifying cell subpopulations in biological fluids aids in diagnosis and understanding of the mechanisms of injury. Although much has been learned from cerebrospinal fluid (CSF) flow cytometry in neuroimmunological disorders such as multiple sclerosis (MS), previous studies did not contain enough healthy donors (HD) to derive age- and gender-related normative data and sufficient heterogeneity of other inflammatory neurological diseases (OIND) controls to identify MS specific changes.

The goals of this blinded, training and validation study of MS patients and embedded controls, representing 1240 prospectively-acquired paired CSF/blood samples from 588 subjects was: 1. To define physiological age/gender-related changes in CSF cells; 2. To define/validate cellular abnormalities in blood and CSF of untreated MS through disease duration (DD) and determine which are MS-specific; 3. To compare effect(s) of low-efficacy (i.e., interferon-beta [IFN-beta] and glatiramer acetate [GA]) and high-efficacy drugs (i.e., natalizumab, daclizumab and ocrelizumab) on MS-related cellular abnormalities using propensity score matching.

Physiological gender differences are less pronounced in the CSF compared to blood, while age- related changes suggest decreased immunosurveillance of CNS by activated, HLA-DR+ T cells associated with natural aging. Results from patient samples support concept of MS being immunologically single disease evolving in time. Initially, peripherally activated innate and adaptive immune cells migrate into CSF to form MS lesions. With progression, T cells (CD8+ > CD4+), NK cells and myeloid dendritic cells are depleted from blood as they continue to accumulate, together with B cells, in the CSF and migrate to CNS tissue forming compartmentalized inflammation. All MS drugs inhibit non-physiological accumulation of immune cells in the CSF. While low efficacy drugs tend to normalize it, high efficacy drugs overshoot some aspects of CSF physiology suggesting impairment of CNS immunosurveillance. Comparable inhibition of MS-related CSF abnormalities advocates changes within CNS parenchyma responsible for differences in drug’s efficacy on MS disability progression.

Video summarizing all results may become useful educational tool.

## 1 Introduction

Neuroimmunological diseases of the central nervous system (CNS) are expanding group of immune-mediated conditions that affect brain and spinal cord. Although last decade brought exciting advances in antibody (Ab)-based diagnostic testing for some of these conditions (1,2), the diagnosis of others is based on combination of clinical suspicion, CNS imaging and evaluation of cerebrospinal fluid (CSF). Examining cellular compositions of CSF may aid in diagnostic process. Because cytospin provides limited information and low cellularity in non-infectious neuroimmunological disorders prevents use of CSF cell blocks, cellular abnormalities in neuroimmunological diseases, including multiple sclerosis (MS) are investigated by flow cytometry. Nevertheless, CSF flow cytometry remains a research test, not performed routinely by clinical laboratories.

In 2014 we published our initial experience (i.e., first 221 subjects) with prospective blood and CSF immunophenotyping of untreated subjects who entered natural history protocol: “Comprehensive Multimodal Analysis of patients with Neuroimmunological Diseases of the CNS”; Clinicaltrials.gov identifier NCT00794352 (3). This study identified new and validated previously published changes in immune compositions of blood versus CSF, as well as changes between different neurological diseases.

However, our 2014 study, as well as most MS blood/CSF immunophenotyping studies published thus far, have following limitations: 1. While some studies did not even consider effects of age and gender (4,5), others, including ours, used age and gender as covariates in the statistical models. However, age and gender are also associated with important aspects of MS phenotype: e.g., females are over-represented among MS patients, especially in relapsing-remitting (RRMS) stage and may have better prognosis than males. Age is linked to immune senescence and progressive accumulation of disability in MS. Therefore, simple statistical adjustments for age/gender effects may mask changes relevant to MS. To discern disease specific effects, more appropriate analysis is to define effects of gender and age in healthy donors (HD) and then subtract physiological effects from patient data. 2. To facilitate conceptual thinking, previous studies either studied single MS clinical phenotype (6,7), or divided MS into traditional RRMS and two progressive (PMS) groups: secondary progressive (SPMS) and primary progressive (PPMS) (8,9). Instead, analyzing changes across all MS patients in relationship to time (e.g., disease duration [DD]), may support or refute a notion that RRMS and PMS represent different stages of the identical, continuously evolving disease. 3. Studies that investigated effect of drugs on blood/CSF cellular compositions usually investigated only one drug at a time and rarely included HDs (10-12). This precludes comparison between drugs and determination whether a drug only inhibits or fully normalizes the studied parameter or, alternatively, exerts effects exceeding physiological ranges. The latter may signify impairment of immune functions and foresee possibility of adverse events.

Based on the value of sensitive enumeration of immune cells in the CSF to aid the diagnosis of neuroimmunological diseases and guide therapeutic decisions, we continue to prospectively acquire blood/CSF flow cytometry-based immunophenotyping data on all subjects in NCT00794352 natural history protocol. This paper addresses the afore-mentioned limitations, while also benefitting from recent advances in data analyses such as propensity score matching.

## 2 Materials and Methods

### 2.1 Patients and Protocol

This study was conducted at the National Institute of Allergy and Infectious Diseases (NIAID) of the National Institutes of Health (NIH) with approval by the Institutional Review Board. All subjects provided written informed consent and were recruited into “Comprehensive Multimodal Analysis of patients with Neuroimmunological Diseases of the CNS”; Clinicaltrials.gov identifier NCT00794352. Samples were collected prospectively, between February 2011 and August of 2019, representing a total of 1240 paired samples of blood and CSF collected from 588 individuals.

Before sample collection, all participants underwent a comprehensive diagnostic process, including neurological examination transcribed to iPad-based app NeurEx (13), which automatically calculates Expanded Disability Status Scale (EDSS; ordinal scale from 0-10) (14) and Combinatorial Weight-Adjusted Disability Score (CombiWISE; continuous scale from 0-100) (15). All subjects had MRI imaging of the brain and upper cervical spinal cord. Clinical and imaging data were transcribed to research database and final diagnosis, together with the rating of diagnostic certainty (as “Definite”, “Most likely” and “Remains Undiagnosed”) was determined and adjusted based on longitudinal follow-up. MS diagnosis was based on 2010 McDonald diagnostic criteria (16) and, after 2017, based on 2017 modifications (17). During longitudinal follow-up initiation and termination dates of any immunomodulatory disease-modifying treatment (DMT) were recorded in the database.

Other inflammatory neurological disorders (OIND) included: cryptococcal meningitis (33/90 patients), Lyme disease with CNS involvement (10/90), sarcoid (7/90), Susac’s syndrome (5/90), neuromyelitis optica (4/90), vasculitis (2/90), Sjogren’s syndrome (2/90), and others (27/90). Non-inflammatory disorders (NIND) included Lyme disease with no CNS involvement (43/80), vascular/ischemic disorders (7/80), migraines (6/80), systemic cryptococcosis without CNS involvement (6/80), and others (19/80).

Drug therapy status was adjudicated based on following criteria: 1. To be classified as untreated a patient was either never treated or had to be off a low efficacy drug for at least 90 days; off a high efficacy drug for at least 180 days and off steroids for at least 30 days. The dichotomization of MS drugs to low versus high efficacy was based on published meta-analysis (18). 2. Treated patient had to be on a drug for a minimum of 90 days. Drug effect(s) were analyzed only when research database contained a minimum of 10 treated patients/drug that fulfilled entry criteria.

### 2.2 Sample Collection, Processing, and Flow Cytometry

Biological samples were collected between 9am-noon, assigned alphanumeric code that blinded the investigators to the diagnosis, treatment status, clinical and imaging data, and processed according to standard operating procedures (SOPs). Briefly, ~25 mL of CSF was collected, transported on ice, and processed within 30 minutes of collection. During processing, CSF cells were transferred from collection tubes into two 15mL centrifuge tubes (Sarstedt, REF: 62554) and spun for 10 minutes at 1200rpm at 4 ^o^C. The pelleted CSF cells were combined and resuspended in X-VIVO™ 15 (Lonza, REF: 04-418Q) on ice. For whole blood, the samples were transferred from collection tubes into BD Vacutainer® CPT^TM^ tubes (BD Biosciences, REF: 362753) to isolate mononuclear cells from whole blood. CPT^TM^ tubes were spun according to manufacturer instructions for 30 minutes at 1800g at room temperature, and the cell portion was transferred into 15 mL centrifuge tubes. Blood cells were washed twice in phosphate buffered saline (PBS) (Gibco, REF: 100100-031) and resuspended in X-VIVO^TM^ 15 on ice. CSF cells were concentrated at 100 cells/μl and cells from blood were concentrated at 2000 cells/μl. Concentrated cells were counted by staining with trypan blue and using hemocytometer and stored on ice prior to staining.

Minimum of 2,000 CSF cells and 200,000 blood cells were transferred into a 96-well plate, and Fc receptors on cells were blocked using 2% intravenous human immunoglobulin (Gamunex-C, REF: NDC 13533-800-20) on ice for 5 minutes. The cells were pelleted for 5 minutes at 1200rpm at 4°C and supernatant discarded. A 12-color Ab panel (Supplementary Table 1) was optimized in preliminary experiments to assure saturating concentrations of each antibody. Antibody-mixture was added to the pelleted cells and mixed gently with pipette, then incubated in dark on ice for 30 minutes, according to published protocol (3). After staining, cells were washed twice, pelleted by centrifuging at 1200rpm for 5 minutes at 4°C, then resuspended in 200μl of fluorescence-activated cell sorting (FACS) buffer (1g of sodium azide (Sigma, REF: 26682-2-8), 10mL of fetal bovine serum (Gemini, REF:100-106), and 1L of PBS (Gibco, REF: 100100-031)). After resuspending, the cells were immediately run in a BD Bioscience LSR II flow cytometer with High Throughput Sampler. Cell populations were gated prospectively using BD FACSDiva software, based on known distributions of cell surface markers for markers of bimodal distribution and based on isotype control for other markers. Gating strategy is described in Figure 1. Gating was quality controlled weekly and adjusted if gating mistakes were identified before uploading results to research database and locking them from further modifications. After defining entry criteria for this study, all eligible prospectively gated results were exported for retrospective analyses.

**Figure 1:**
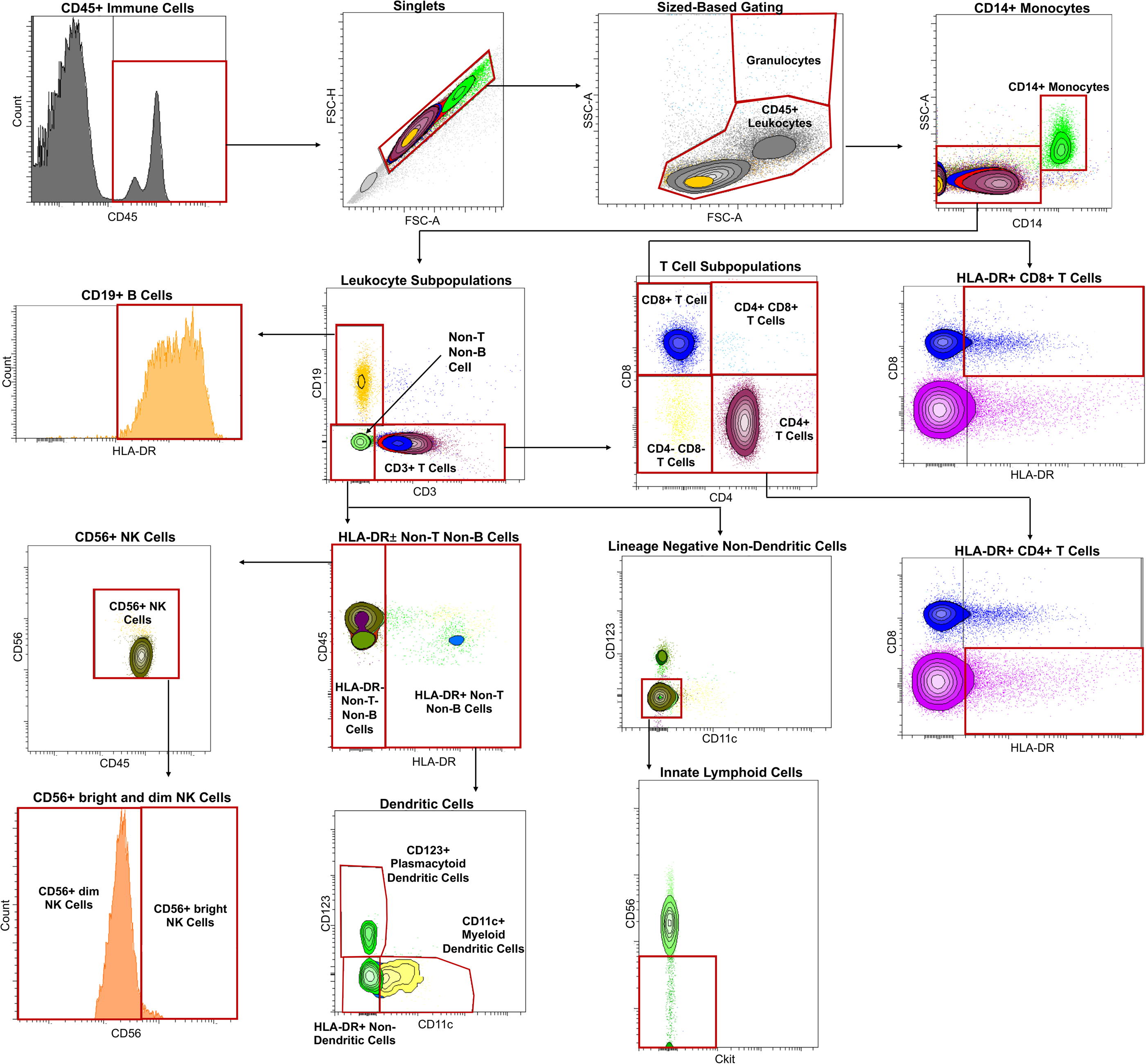
Immune cells were first classified by CD45 expression with histogram gating, followed by gating into singlets with size-based gating (forward scatter area [FSC-A] vs. forward scatter height [FSC-H]). Granulocytes were distinguished from CD45+ leukocytes with size-based gating as well (FSC-A vs. side scatter [SSC]-A). Monocytes were distinguished CD45+ leukocytes using CD14 expression. Leukocyte subpopulations were then identified by subtracting CD14+ monocytes from CD45+ leukocytes and categorized by lack or expression of CD19 or CD3 into adaptive and innate immunity. CD19 expressing cells were confirmed to be CD19+ B cells by gating using of HLA-DR. T cells were identified using CD3 expression, with further division into CD4 expressing T cells and CD8 expressing T cells. Activated or HLA-DR expressing CD4+ T cells and CD8+ T cells were also identified in the same gate by placing HLA-DR in the x-axis and CD8 in the y-axis. Cells lacking CD19 and CD3 expression were categorized into innate immunity and further subdivided by HLA-DR expression. HLA-DR expressing cells as part of innate immunity were divided into dendritic cell populations, with myeloid dendritic cells identified using CD11c and plasmacytoid dendritic cells using CD123. HLA-DR negative cells were on the other hand identified into NK cell populations by using CD56. CD56+ NK cells were then divided into CD56^dim^ and CD56^bright^ cells using CD56 expression as a guide. Innate lymphoid cells were identified from cells lacking CD3 vs. CD19, CD11c vs. CD123, and Ckit vs. CD56.

### 2.3 Logarithmic Transformation and Outlier Analysis

All analysis was performed in R Studio statistical software (19).

Appropriate cell ratios (number of cells in a population/number of cells in another population), cell proportions (number of cells in a population/number of CD45+ leukocytes), and absolute numbers (number of cells/ml) were calculated, and log transformed. Outlier analysis was completed independently in each diagnostic category by excluding any data points in a cell ratio, cell proportion, or absolute number that was outside first quarter (Q1) − 3*interquartile range (IRQ) or third quarter (Q3) + 3*IQR.

### 2.4 Age/Gender Adjustments

All cell proportions and absolute numbers were correlated with age separately in CSF, blood, and as CSF/blood ratios using Spearman correlation in HD cohort. Features that demonstrated significant correlation defined as *p*-value ≤ 0.05 adjusted for multiple comparisons using false discovery rate (FDR) were subsequently adjusted in patient samples using linear regressions derived from HD cohort: specifically, the measured values were re-calculated as residuals from HD linear regressions as previously described (20).

The same was performed for analysis of gender differences using unpaired t-test, where physiological gender effects that remained significant after adjustment for multiple comparisons were subtracted from one gender category as group medians. All features that were found to be affected by age or gender can be found in Tables 1 and 2 with examples of adjustments in Figure 2. The R-code for age/gender adjustments can be found in GitHub, with link in Data Availability section.

**Table 1:** Immunophenotyping features were correlated in 48 HDs with age in blood, CSF, and CSF/blood ratios. Correlation coefficient and spearman *p*-values were calculated after adjustment for multiple comparisons.

| Blood (n=48) |  |  | CSF (n=48) |  |  | CSF/Blood (n=48) |  |  |
| --- | --- | --- | --- | --- | --- | --- | --- | --- |
| Feature | Spearman Rho | p-value | Feature | Spearman Rho | p-value | Feature | Spearman Rho | p-value |
| HLA-DR+ CD4+ T-Cell/CD45+ Leukocyte Proportion | 0.389 | 0.007 | CD4- CD8- T Cell/CD45+ Leukocyte Proportion | -0.401 | 0.007 | CD19+ B-Cell/CD14+ Monocyte Ratio | -0.440 | 0.003 |
| CD11c+ Myeloid Dendritic Cell/CD123+ Plasmacytoid Dendritic Cell Ratio | 0.387 | 0.007 | CD19+ B-Cell/CD14+ Monocyte Ratio | -0.385 | 0.009 | Innate Lymphoid Cell Abs | -0.390 | 0.021 |
| CD123+ Plasmacytoid Dendritic Cell/HLA-DR+ non-T non-B Cell Ratio | -0.387 | 0.008 | CD19+ B-Cell/CD45+ Leukocyte Proportion | -0.355 | 0.014 | HLA-DR+ CD8+ T-Cell Abs | -0.338 | 0.022 |
| CD4+ T-Cell Abs | 0.386 | 0.009 | CD4+ T-Cell/CD3+ T-Cell Ratio | 0.337 | 0.024 | HLA-DR+ CD4+ T-Cell Abs | -0.337 | 0.024 |
| CD56+ NK Cell Abs | 0.349 | 0.018 |  |  |  |  |  |  |
| CD4+ T-Cell/CD45+ Leukocyte Ratio | 0.336 | 0.021 |  |  |  |  |  |  |
| CD56+ dim NK Cell Abs | 0.337 | 0.023 |  |  |  |  |  |  |
| HLA-DR+ CD8+ T-Cell Abs | 0.319 | 0.031 |  |  |  |  |  |  |
| CD3+ T-Cell/CD45+ Leukocyte Proportion | 0.320 | 0.035 |  |  |  |  |  |  |
| CD3+ T-Cell Abs | 0.309 | 0.037 |  |  |  |  |  |  |
| CD4+ T-Cell/CD3+ T-Cell Ratio | 0.305 | 0.038 |  |  |  |  |  |  |
| HLA-DR+ CD4+ T-Cell Abs | 0.302 | 0.038 |  |  |  |  |  |  |
| CD56+ bright NK Cell Abs | 0.305 | 0.040 |  |  |  |  |  |  |
| HLA-DR+ CD8+ T-Cell/CD8+ Cell Ratio | 0.296 | 0.044 |  |  |  |  |  |  |
| CD11c+ Myeloid Dendritic Cell Abs | 0.292 | 0.049 |  |  |  |  |  |  |

**Table 2:** T-test with adjustment for multiple comparisons was performed in CSF and blood to identify gender differences in immunophenotyping features. *p*-values were calculated along with male and female mean values.

| Blood (n=48) |  |  |  | CSF (n=48) |  |  |  |
| --- | --- | --- | --- | --- | --- | --- | --- |
| Clinical Marker: | *p*-value | Male Mean (n=24): | Female Mean (n=24): | Clinical Marker: | *p*-value | Male Mean (n=24): | Female Mean (n=24): |
| CD4+ T-Cell/CD3+ T-Cell Ratio | 0.002 | -0.557 | -0.425 | CD56+ NK Cell/CD45+ Leukocyte Proportion | 0.011 | -4.190 | -3.795 |
| CD4+ T-Cell/CD8+ T-Cell Ratio | 0.008 | 0.614 | 0.918 | CD56+ bright NK Cell/CD45+ Leukocyte Proportion | 0.021 | -4.827 | -4.443 |
| CD19+ B-Cell/CD45+ Leukocyte Proportion | 0.010 | -2.546 | -2.283 | CD4+ T-Cell/CD45+ Leukocyte Proportion | 0.024 | -0.764 | -0.620 |
| CD19+ B-Cell/CD14+ Monocyte Ratio | 0.011 | -0.345 | 0.202 | CD4+ T-Cell/CD8+ T-Cell Ratio | 0.041 | 1.111 | 1.343 |
| HLA-DR+ CD8+ T-Cell/CD45+ Leukocyte Proportion | 0.012 | -3.259 | -3.918 |  |  |  |  |
| CD8+ T-Cell/CD3+ T-Cell Ratio | 0.015 | -1.171 | -1.384 |  |  |  |  |
| HLA-DR+ CD4+ T-Cell/CD4+ T-Cell Ratio | 0.029 | -2.428 | -2.786 |  |  |  |  |
| CD56+ dim NK Cell/CD56+ bright NK Cell Ratio | 0.033 | 2.948 | 2.546 |  |  |  |  |
| CD4- CD8- Cell/CD45+ Leukocyte Ratio | 0.040 | -3.128 | -3.455 |  |  |  |  |
| HLA-DR+ CD8+ T-Cell/CD8+ T-Cell Ratio | 0.044 | -1.561 | -2.001 |  |  |  |  |

**Figure 2:**
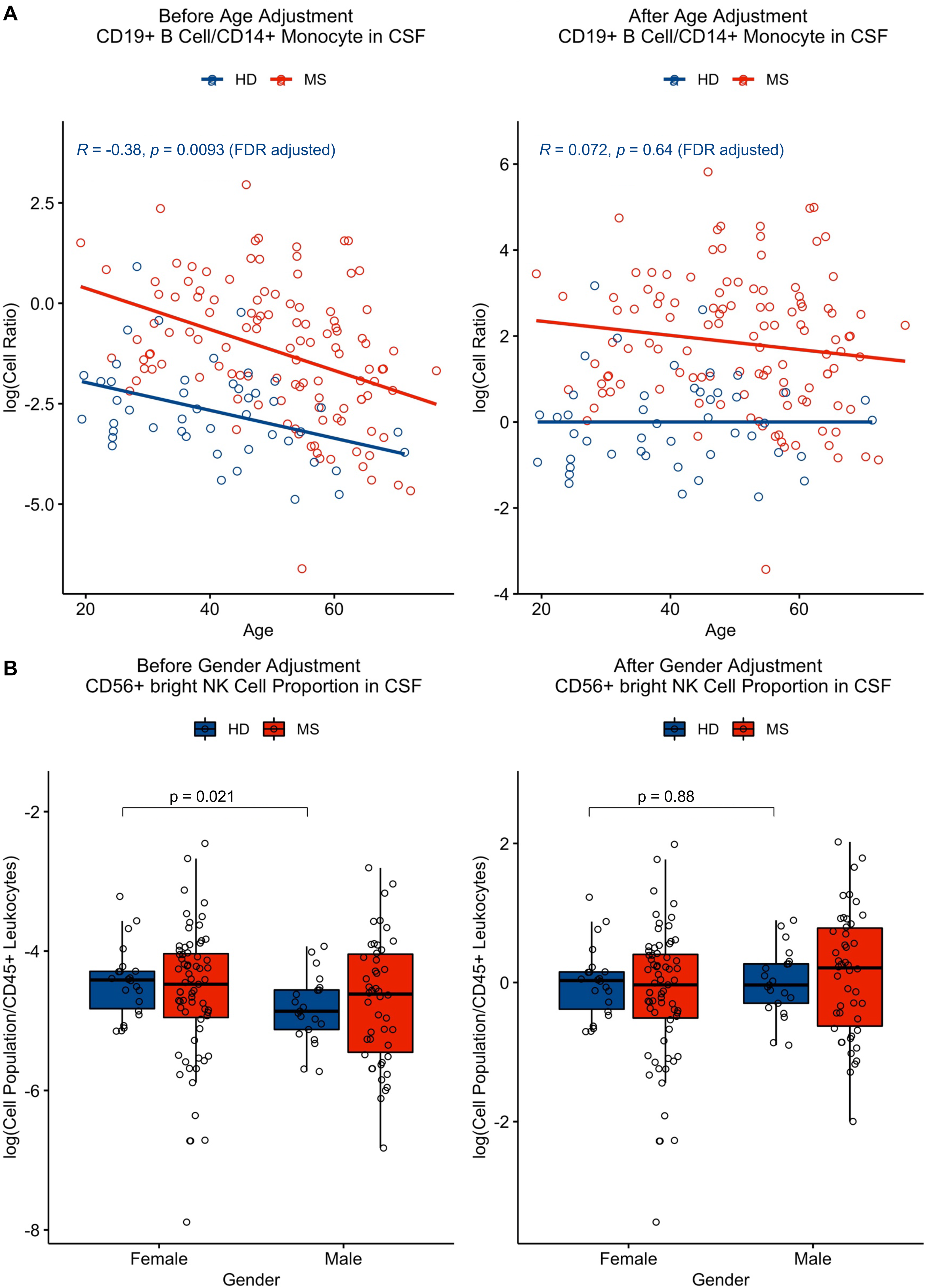
(A) Features that correlated significantly (*p*-value ≤ 0.05) with age in HDs (in blue) were adjusted using linear regression. This adjustment was applied to all patient cohorts, including the MS patient cohort (as shown in red). (B) Features that had significant gender differences (*p*- value ≤ 0.05) were adjusted as well, with application of this adjustment to all cohorts by subtracting one gender category as group medians.

### 2.5 Training and Validation Cohorts

To find reproducible changes in age/gender-adjusted flow cytometry parameters between MS patients and HDs, while limiting number of comparisons, we divided untreated MS patients into training and validation cohorts with a 50/50 split, accounting for comparable distribution of age, gender, disability, and MS subtypes. Unpaired t-test analyzed differences between HDs and MS patients in the training cohort first. Significantly different markers (*p*-value ≤ 0.05, FDR adjusted) from the training cohort were assessed in the validation cohort. Only statistically significant and reproducible features are displayed and discussed here. All validated markers are shown in Table 3. Demographic details of untreated MS patients in training and validation cohorts are described in Supplementary Table 2.

**Table 3:**
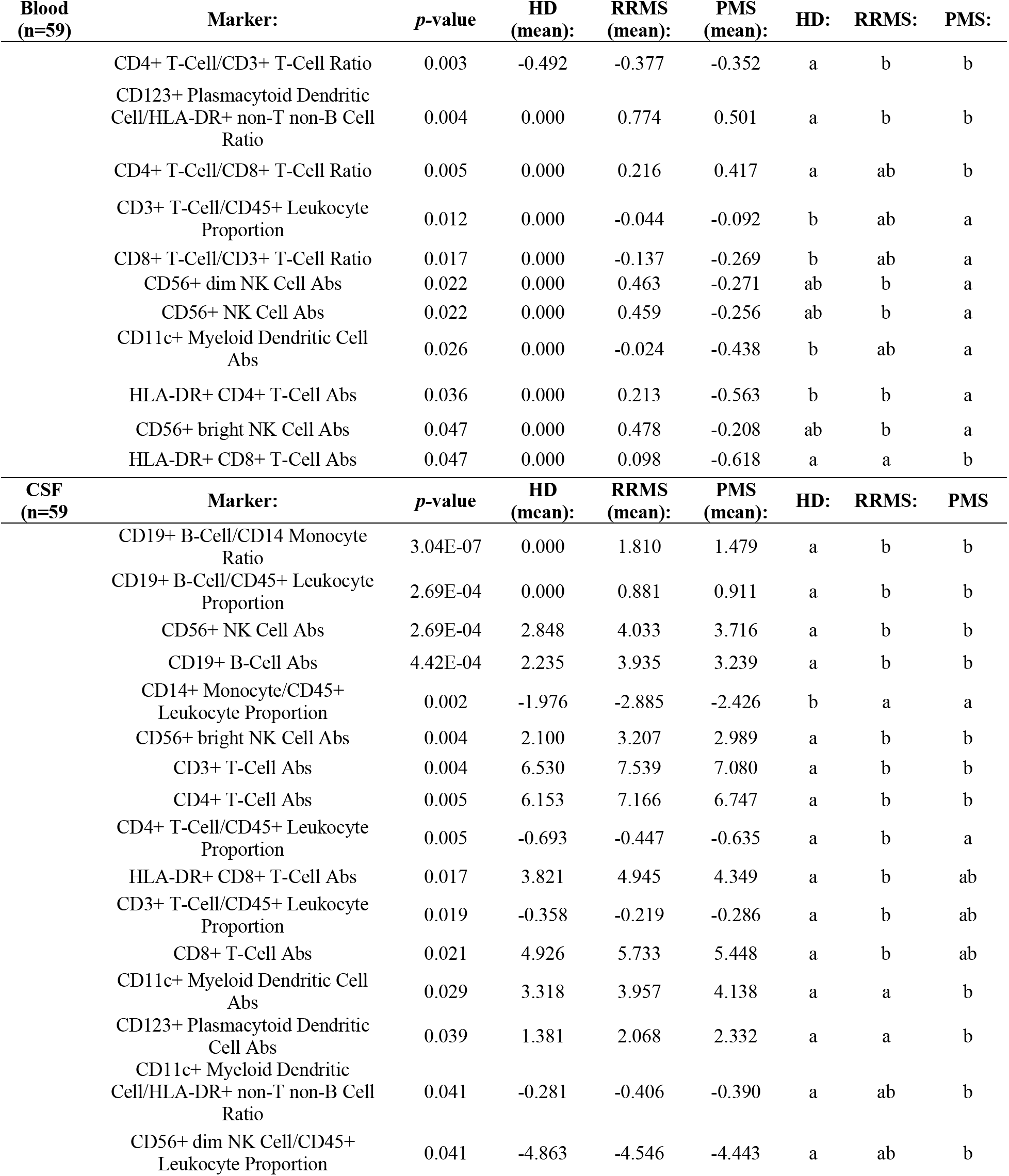
Features that were statistically different from each other between HDs and RRMS and/or PMS in training cohort were then validated in an independent validation cohort and features significant in validation cohort described here. For each feature, *p*-value adjusted for multiple comparisons is shown, along with mean values of HDs, RRMS, and PMS cohorts, and letters describing which cohorts are significantly different from each other. Cohorts with same letters are insignificantly different from each other in the given feature, meanwhile cohorts with different letters are significantly different from each other in that feature.

### 2.6 Propensity Score Matching to Investigate Effects of MS Therapies

Because our natural history cohort included limited number of MS patients treated with specific drugs, there was high likelihood of unequal distribution of demographic data between smaller drug-treated cohorts and the large untreated cohort. Thus, we employed propensity score matching, where each treated MS cohort was matched 1:3 to highly comparable cohort of untreated MS patients in terms of age, gender, and level of disability. Differences between treated and propensity score matched untreated MS patients were evaluated using unpaired t-test with adjustment for multiple comparisons. This data was supplemented with longitudinal paired data (i.e., before and after specific therapy for all subjects that had such follow-up CSF in NSD database) to demonstrate agreement between propensity score matched group results and intra-individual before-after treatment results. Due to limited sample sizes, no formal statistics was applied to these before/after comparisons.

Figure 3 shows propensity score matched age, gender, and disability scales for matched untreated MS and treated MS cohorts. Meanwhile Table 4 describes all patient categories and their demographics.

**Figure 3:**
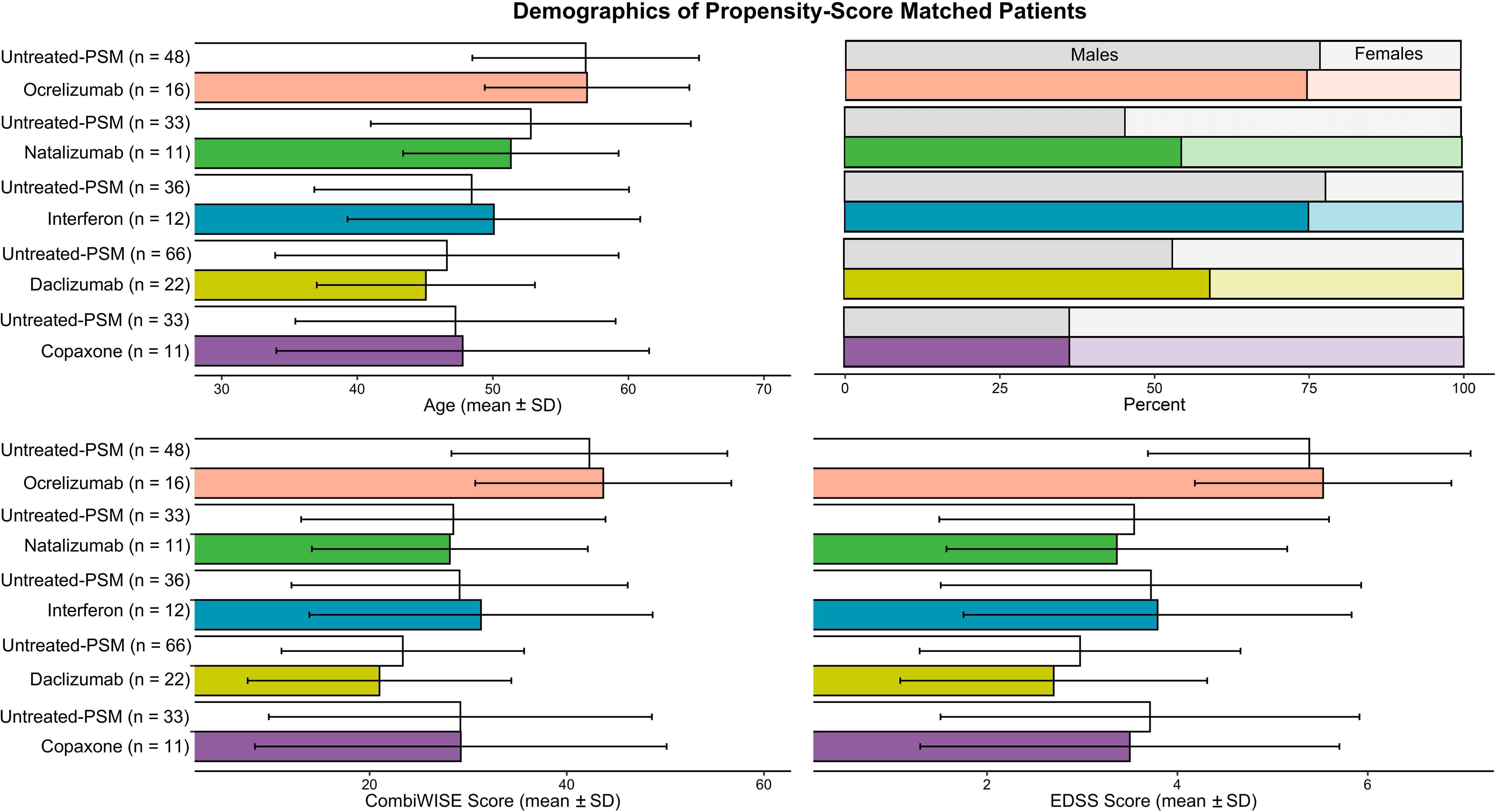
To compare features in untreated and treated MS patients, propensity score matching was performed to allow for appropriate comparisons in features while accounting for differences in age, gender, and disability. Each matched untreated and treated patient group means and ± SD were graphed. For each drug-treated MS patient group, the algorithm picked out from 118 untreated MS patients a cohort of untreated MS patients that is three times larger than the treated group, while also making sure age, gender, and CombiWISE were as close to the treated group as possible.

**Table 4:**
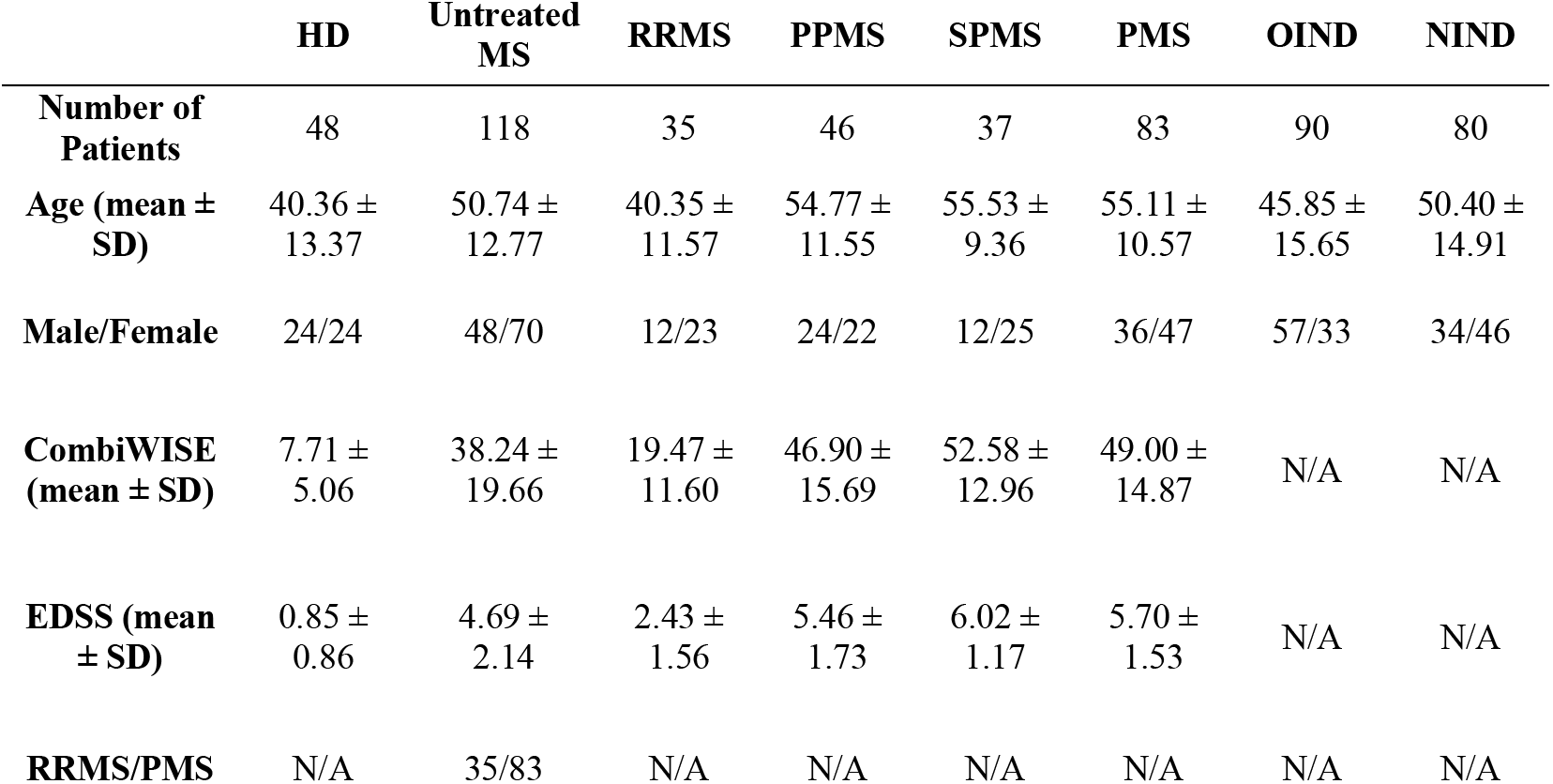
Demographics of untreated patient cohorts are described here, specifically number of patients, age, gender, disability, and MS subtype.

|  | HD | Untreated MS | RRMS | PPMS | SPMS | PMS | OIND | NIND |
| --- | --- | --- | --- | --- | --- | --- | --- | --- |
| **Number of Patients** | 48 | 118 | 35 | 46 | 37 | 83 | 90 | 80 |
| **Age (mean ± SD)** | 40.36 ± 13.37 | 50.74 ± 12.77 | 40.35 ± 11.57 | 54.77 ± 11.55 | 55.53 ± 9.36 | 55.11 ± 10.57 | 45.85 ± 15.65 | 50.40 ± 14.91 |
| **Male/Female** | 24/24 | 48/70 | 12/23 | 24/22 | 12/25 | 36/47 | 57/33 | 34/46 |
| **CombiWISE (mean ± SD)** | 7.71 ± 5.06 | 38.24 ± 19.66 | 19.47 ± 11.60 | 46.90 ± 15.69 | 52.58 ± 12.96 | 49.00 ± 14.87 | N/A | N/A |
| **EDSS (mean ± SD)** | 0.85 ± 0.86 | 4.69 ± 2.14 | 2.43 ± 1.56 | 5.46 ± 1.73 | 6.02 ± 1.17 | 5.70 ± 1.53 | N/A | N/A |
| **RRMS/PMS** | N/A | 35/83 | N/A | N/A | N/A | N/A | N/A | N/A |

## 3 Results

Because of the quantity of statistically significant results and the complexity of interpretations that integrate paired blood and CSF samples, we developed schematic representations of each results section (Supplementary Presentation 1) and integrated these into a single animated video (Supplementary Video 1).

### 3.1 Physiological Effects of Age and Gender on Cellular Subpopulations of Blood and CSF

#### 3.1.1 Age

Within HDs, we observed (Table 1; Supplementary Presentation 1, Slide 2) the following changes in cellular subpopulations with age in the blood: Both CD3+ T cells and CD4+ T cells correlated positively with age as an absolute number and as a proportion. Multiple HLA-DR+ T-cell populations also correlated positively with age, including both HLA-DR+ CD4+ T cell and HLA-DR+ CD8+ T cell absolute numbers and proportions. All NK cell absolute numbers correlated positively with age, including CD56^bright^ and CD56^dim^ NK subsets. Of dendritic cell populations, the absolute number of CD11c+ myeloid dendritic cells (myDC) correlated positively with age along with ratio of CD11c+ myeloid dendritic cells to CD123+ plasmacytoid dendritic cells (plDC). CD123+ plDC correlated negatively with age as a ratio to HLA-DR+ non-T non-B cells.

In the CSF we observed a positive correlation of age with CD4+ T cell/CD3+ T cell ratio. Both proportions of CD4- CD8- T cell and CD19+ B cells correlated negatively with age in the CSF, along with CD19+ B-cell/CD14+ monocyte cell ratio.

Several markers as CSF/blood ratios also correlated negatively with age, including both absolute numbers of HLA-DR+ CD4+ T cells and HLA-DR+ CD8+ T cells. The absolute number of innate lymphoid cells and cell ratio of CD19+ B cells/CD14+ monocytes correlate negatively with age.

#### 3.1.2 Gender

Within HDs, we observed (Table 2; Supplementary Presentation 1, Slide 3) following gender-specific differences in cellular subpopulations in the blood: Males had higher proportions of CD4- CD8- T cells, HLA-DR+ CD4+ T cell, HLA-DR+ CD8+ T cells, and CD8+ T cells. Males had also increased CD56^dim^/CD56^bright^ NK cell ratio. Females, on the other hand had a higher proportion of CD4+ T cells, CD19+ B cells, CD4/CD8 T cell ratio, and CD19+ B cells/CD14+ monocytes ratio.

In the CSF, following markers were higher in females compared to males: proportion of CD4+ T cells, CD4/CD8 T cell ratio, and proportion of both CD56+ NK cells and CD56^bright^ NK cells.

We did not observe any gender differences in CSF/blood ratios.

### 3.2 Comparison of Cellular Changes Between HDs and Untreated MS Patients

#### 3.2.1 Relapsing-Remitting MS (RRMS)

In RRMS blood (Figure 4; Supplementary Presentation 1, Slide 4), the proportion of CD4+ T cells (out of all CD3+ T cells) was significantly elevated in comparison to HDs. This increase was MS-specific and can’t be observed in OIND and NIND controls. Of the dendritic cell populations in RRMS, the proportion of CD123+ plDC was increased.

**Figure 4:**
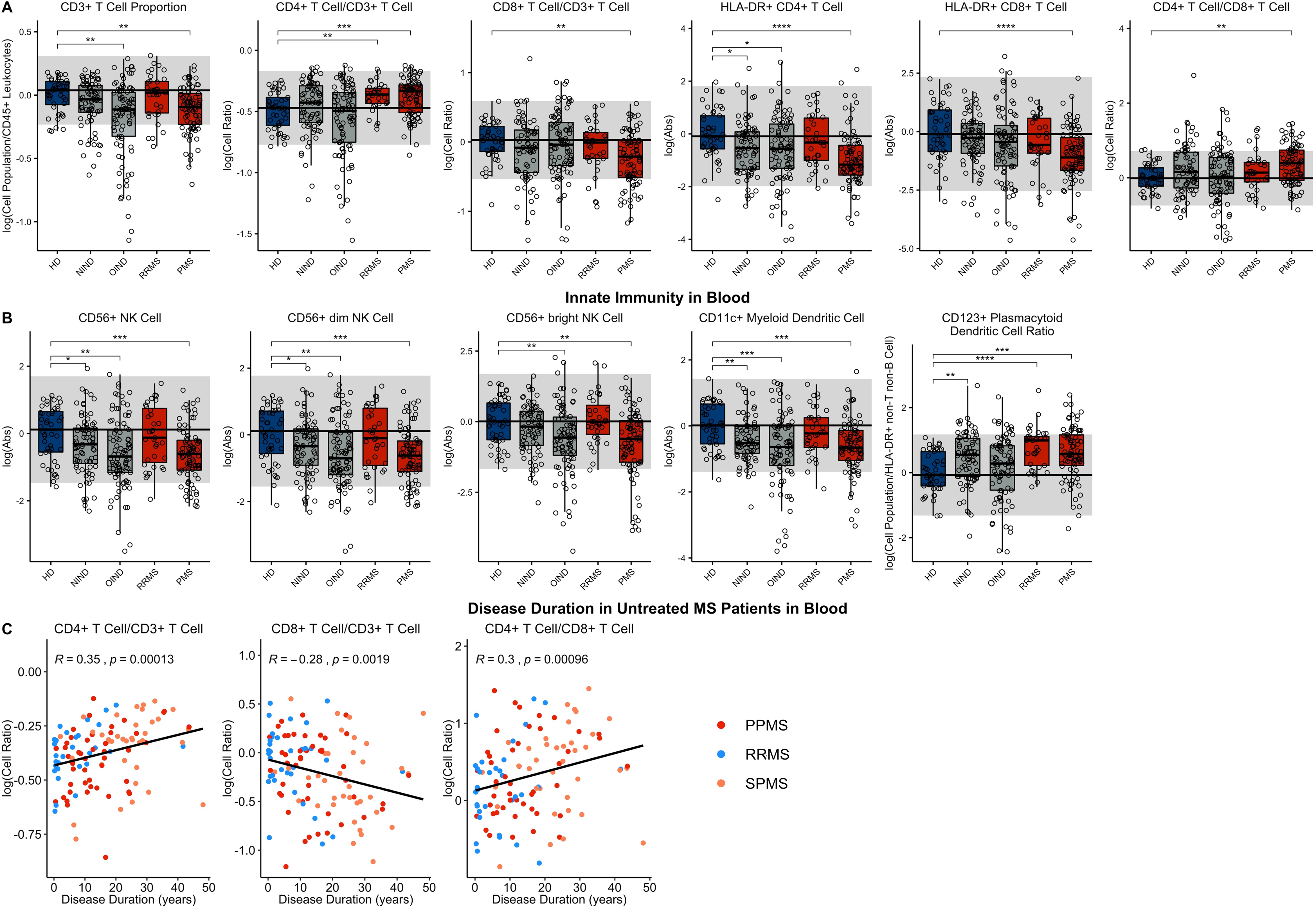
Features that validated in the independent validation cohort in blood during comparison of HDs to RRMS and PMS patients were graphed. HDs, NINDs, OINDs, RRMS, and PMS cohorts for each significant feature were graphed. Unpaired t-test was completed to compare each cohort to HDs with adjustment for multiple comparisons. \**p* < 0.05, **0.01 *<p* < 0.005, *\*\*\*p* < 0.001, \*\*\*\**p* < 0.0001. HD median for each feature was also graphed horizontally and grey shading added representing ± 2 SDs of each feature in HD cohort. (A) Features in adaptive immunity. (B) Features in innate immunity. (C) Features that validated were correlated with disease duration and statistically significant features graphed (*p*-value ≤ 0.05). MS subtypes were color coded to show heterogeneity of cohorts in each significant feature, with blue representing RRMS, red representing PPMS, and orange representing SPMS patients.

In RRMS CSF (Figure 5; Supplementary Presentation 1, Slide 4), both the proportion and absolute number of all CD3+ T cells and CD4+ T cell subset were increased. For CD8+ T cells, only the absolute numbers of CD8+ T cells and their recently activated HLA-DR+ CD8+ T cell subset were increased. The increase in CD4+ T cell proportion was specific to MS. CD19+ B cells were increased as an absolute number and proportion. As reported previously, the proportion of CD14+ monocytes was decreased in the RRMS CSF. The absolute numbers of both NK cell subsets were increased, but proportionally, CD56^dim^ NK cells predominated. Of the dendritic cell populations, the absolute numbers of both DC subsets were increased, but proportionally, CD11c+ myDC were decreased in RRMS compared to HD.

**Figure 5:**
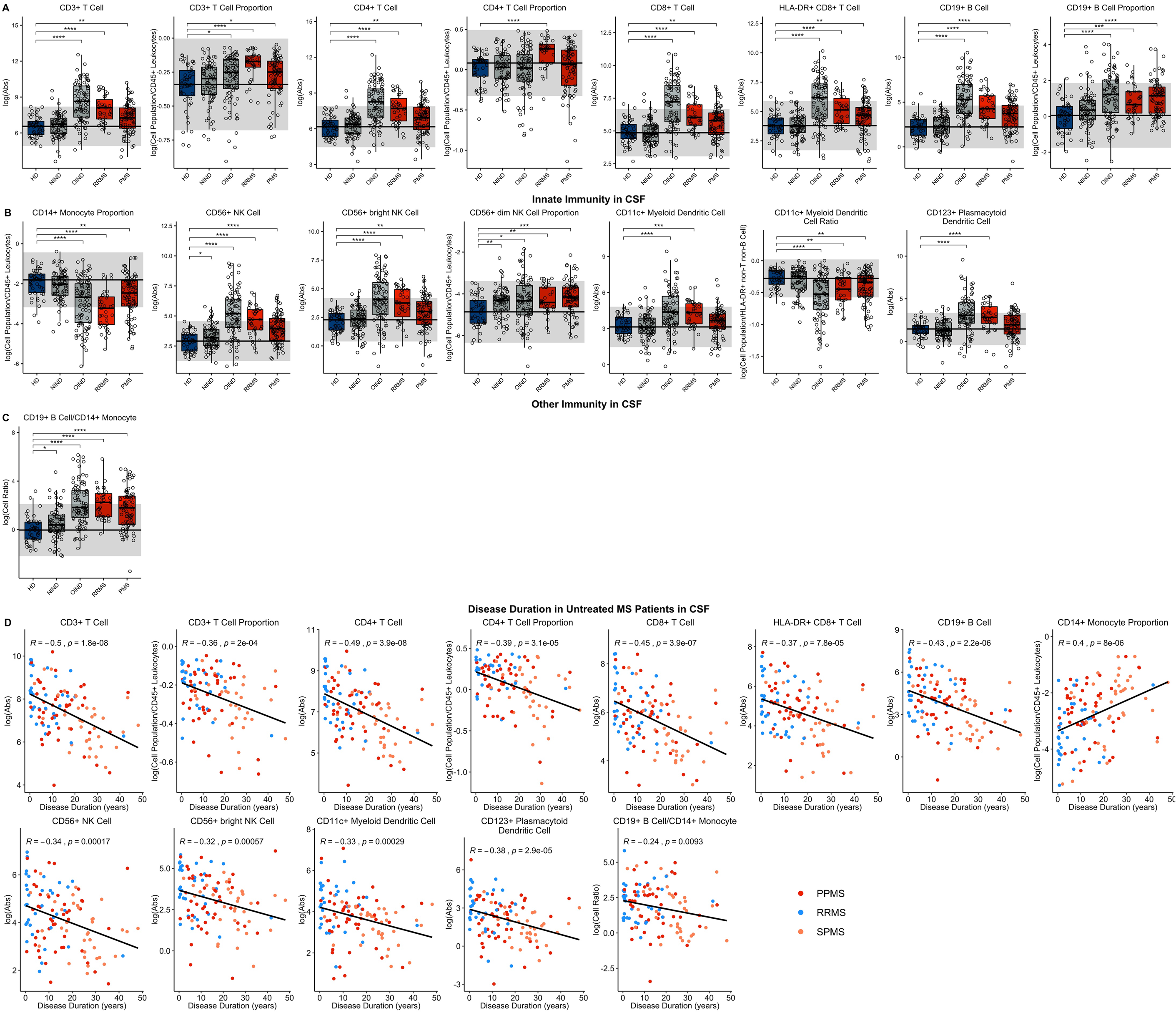
Features that validated in the independent validation cohort in CSF during comparison of HDs to RRMS and PMS patients were graphed. HDs, NINDs, OINDs, RRMS, and PMS cohorts for each significant feature were graphed. Unpaired t-test was completed to compare each cohort to HDs with adjustment for multiple comparisons. \**p* < 0.05, **0.01 *<p* < 0.005, \*\*\**p* < 0.001, \*\*\*\**p* < 0.0001. HD median for each feature was also graphed horizontally and grey shading added representing ± 2 SDs of each feature in HD cohort. (A) Features in adaptive immunity. (B) Features in innate immunity. (C) Features that validated were correlated with disease duration and statistically significant features graphed (*p*-value < 0.05). MS subtypes were color coded to show heterogeneity of cohorts in each significant feature, with blue representing RRMS, red representing PPMS, and orange representing SPMS patients.

We clearly validated previously reported increase in CSF CD19+ B cell/CD14+ monocyte ratio in RRMS.

While to our knowledge all previous studies analyzed separately differences among diagnostic categories in the blood and CSF, we calculate CSF/blood ratios for each subject, to control CSF levels of immune cells for subject-specific levels of immune cell subpopulations in the blood (Supplementary Figure 1). This allowed us to assess migration of the cells from blood to CSF. CSF/blood ratios of all lymphocyte populations of adaptive immunity were higher in RRMS compared to HD. For innate immunity, CSF/blood ratios of all NK cells and both subtypes of DCs were likewise increased in RRMS.

#### 3.2.2 Progressive MS (PMS)

Like in RRMS, PMS blood had increased proportion of CD4+ T cells and increased CD4/CD8 T cell ratio compared to HD (Figure 4; Supplementary Presentation 1, Slide 5). Again, these changes were MS-specific, not observed in other diagnostic groups. However, PMS had decreased blood proportions of all T cells (CD3+), driven by CD8+ T cells subset. The other important change not observed in RRMS was decrease in the absolute numbers of HLA-DR+ CD4+ T cells and HLA-DR CD8+ T cells.

PMS subjects’ blood had also decreased absolute numbers of NK cells – affecting both subtypes - and CD11c+ myDCs. The decrease in myDCs caused significant proportional increase in CD123+ plDCs.

Like we observed in RRMS, PMS CSF had increased absolute numbers of all T cells: CD3+ CD4+, CD8+ and also HLA-DR+ CD8+ (Figure 5; Supplementary Presentation 1, Slide 5). Analogously, CD19+ B cells were increased as an absolute number and proportion.

Also, the innate immune system abnormalities observed in RRMS were reproduced in PMS CSF: decreased proportion of CD14+ monocytes and increased absolute number of NK cells, with CD56^dim^ subset proportionally dominating. Expectedly, CD19+ B cell/CD14+ monocyte ratio was increased in PMS CSF.

The similarity of the CSF phenotype between RRMS and PMS patients suggested identical immune pathophysiology of all MS subtypes. To test this hypothesis, we analyzed correlations of blood and CSF immune populations with MS disease duration (DD). In the blood we observed significant positive associations of CD4+ T cell and CD4/CD8 T cell ratios with MS DD (Figure 4), meanwhile proportion of CD8+ T cells correlated negatively with DD. Untreated MS patients demonstrated highly statistically significant declines in absolute numbers of all immune cells in the CSF, as a function of DD (Figure 5). Only proportion of monocytes increased with DD. The same conclusions were derived from CSF/blood ratios of immune cells (Supplementary Figure 1). The color coding of the 3 MS subtypes in all of these correlation plots provide strong evidence for the uniform immune-cell abnormalities in MS, that evolve as function of DD and not of clinical classification.

### 3.3 Drug-Related Effects on MS Inflammation

Having defined blood and CSF cellular abnormalities of untreated MS (with subtracted effects of natural aging and gender), we next investigated effects of 5 different immunoregulatory treatments of MS using propensity score matching.

#### 3.3.1 Interferon-beta (IFN-beta)

IFN-beta treatment led to highly significant and drug-specific increase in HLA-DR+ T cells, in the blood and CSF. This included all absolute numbers and proportions of HLA-DR+ CD4+ and CD8+ T cells (Figure 6; Supplementary Presentation 1, Slide 6). Also, CD19+ B cell absolute number was elevated in blood of IFN-beta-treated patients compared to matched untreated subjects.

**Figure 6:**
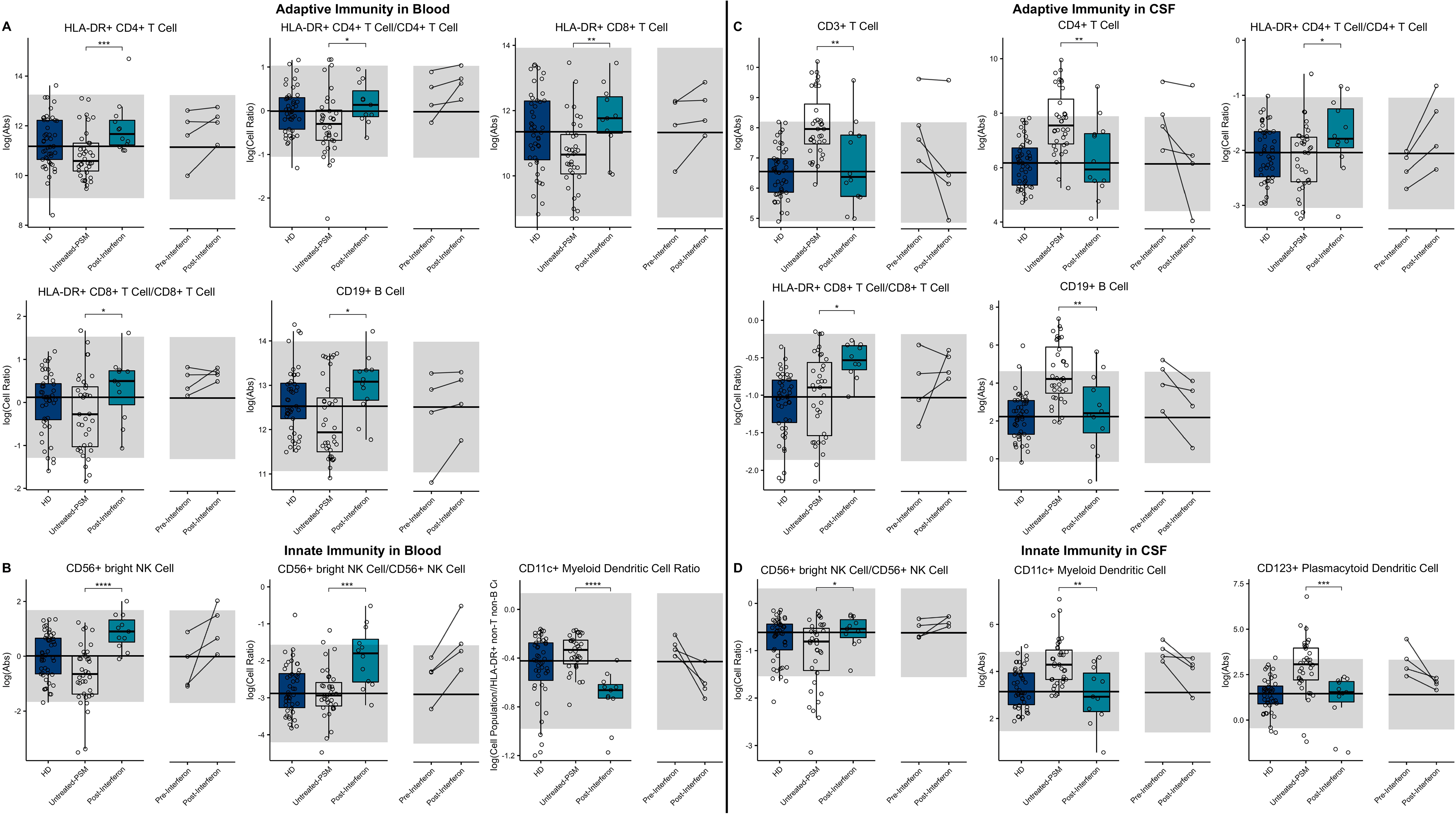
Unpaired t-test with adjustment for multiple comparisons was performed for features during comparison of 36 propensity score matched (PSM) untreated MS patients and 12 interferon-beta treated MS patients in blood and CSF. All significant markers were graphed and supplemented with HD cohort and data from 4 longitudinal patients with paired pre-treatment and post-treatment data. \**p* < 0.05, **0.01 *<p* < 0.005, *\*\*\*p* < 0.001, *\*\*\*\*p* < 0.0001. HD median for each feature was also graphed horizontally and grey shading added representing ± 2 SDs of each feature in HD cohort. (A) Features in adaptive immunity in blood. (B) Features in innate immunity in blood. (C) Features in adaptive immunity in CSF. (D) Features in innate immunity in CSF.

We also reproduced previously published observation that IFN-beta-treatment increased the absolute number of CD56^bright^ NK cells and the ratio of CD56^bright^ to CD56+ NK cells. Finally, we saw highly significant decrease in the proportion of CD11c+ myDCs in the IFN-beta treated subjects.

While the absolute number of T cells, especially CD4+ T cells and CD19+ B cells decreased in the CSF of IFN-beta-treated subjects, the proportions of HLA-DR+ T cells, both CD4+ and CD8+ actually increased, paralleling the same proportional increases of these recently-activated T cells in the blood of IFN-beta-treated subjects. In the innate immunity, the proportional increase of CD56^bright^ NK cells propagated from blood to CSF of IFN-beta-treated subjects. Additionally, IFN-beta decreased absolute numbers of both subsets of DCs.

The investigation of CSF/blood (Supplementary Figure 2) ratios demonstrated that these IFN-beta induced changes on immune cell subpopulations (i.e., expansion of HLA-DR+ T cells and CD56^bright^ NK cells), while observed in both compartments, was much greater in the blood than in the CSF.

#### 3.3.2 Glatiramer Acetate (GA)

We observed that compared to propensity score matched untreated MS, GA-treatment increased absolute numbers and proportions of CD8+ T cells in blood (Figure 7; Supplementary Presentation 1, Slide 7) Surprisingly, GA-treated patients had also higher absolute numbers of immunoregulatory CD56^bright^ NK cells in the blood, while proportion of CD11c+ myDCs was decreased.

**Figure 7:**
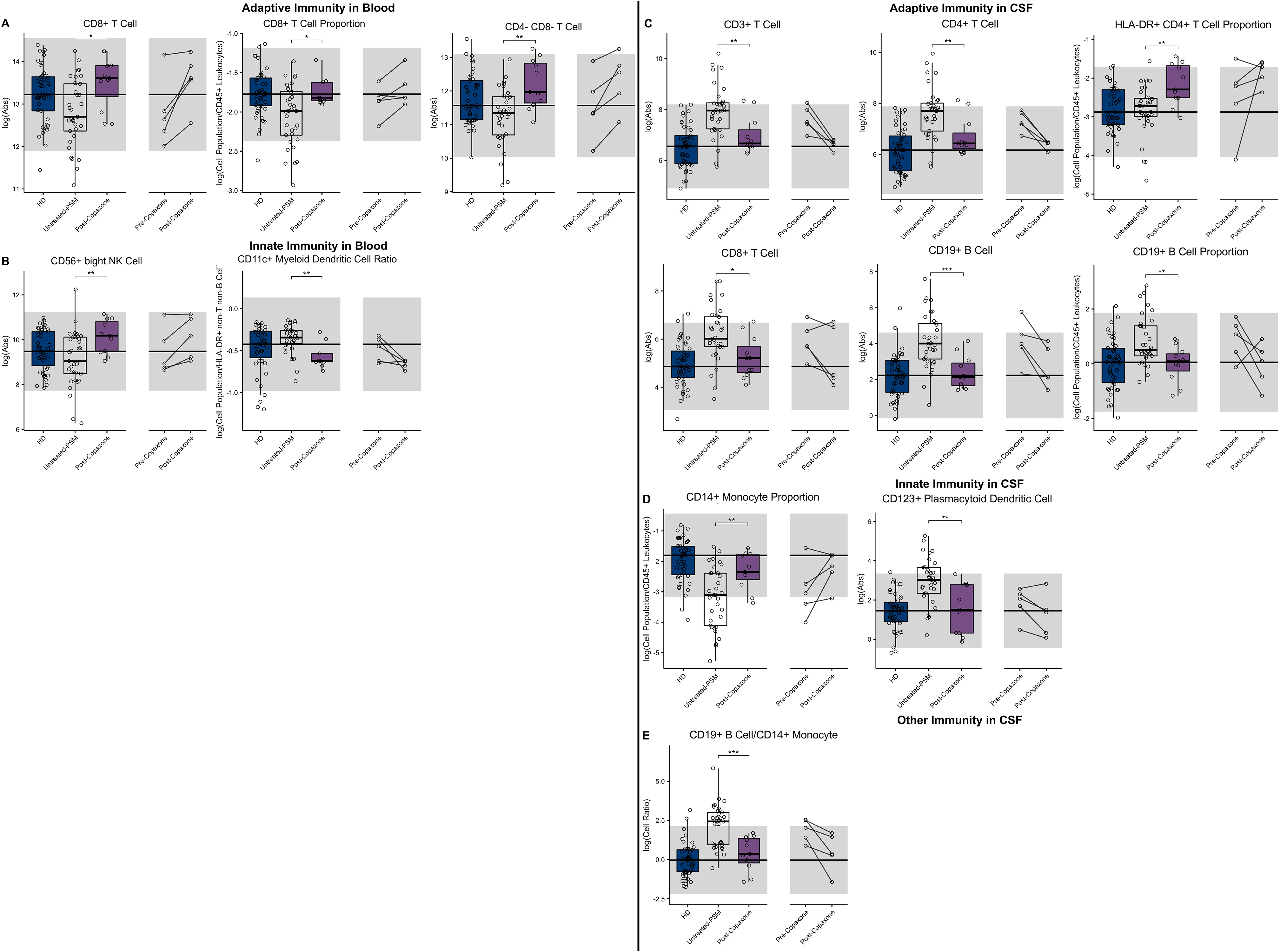
Unpaired t-test with adjustment for multiple comparisons was performed for features during comparison of 33 propensity score matched (PSM) untreated MS patients and 11 glatiramer acetate (GA) treated MS patients in blood and CSF. All significant markers were graphed and supplemented with HD cohort and data from 5 longitudinal patients with paired pre-treatment and post-treatment data. \**p* < 0.05, **0.01 < *p* < 0.005, \*\*\**p* < 0.001, \*\*\*\**p* < 0.0001. HD median for each feature was also graphed horizontally and grey shading added representing ± 2 SDs of each feature in HD cohort. (A) Features in adaptive immunity in blood. (B) Features in innate immunity in blood. (C) Features in adaptive immunity in CSF. (D) Features in innate immunity in CSF. (E) Features in other immunity in CSF.

In the CSF of GA treated MS patients, absolute numbers of all lymphocytes of adaptive immunity (i.e., T and B cells) were significantly decreased compared to untreated MS patients. However, the proportion of HLA-DR+ CD4+ T cells was increased. Within innate immune system cells, GA-treatment elevated CD14+ monocyte proportion in the CSF and normalized B cell/monocyte ratio. GA-treated patients had also normal absolute numbers of plDCs, which were significantly elevated in untreated MS subjects. The examination of CSF/blood ratios (Supplementary Figure 3) only enhanced afore-mentioned observations indicating that the prevalent action of GA is overall normalization of MS-associated CSF inflammation.

#### 3.3.3 Natalizumab

The first high-efficacy MS treatment we examined was natalizumab. Consistent with its mechanism of action (MOA) of inhibiting migration of most immune cells (except granulocytes and to a lesser degree monocytes) from blood to CNS, but also to other organs, we reproduced previously-reported increase in most immune cell subpopulations in the blood of natalizumab-treated patients (Figure 8; Supplementary Presentation 1, Slide 8). Not investigated previously, we observed enrichment of innate lymphoid cells (ILCs), NK cells (CD56^bright^>CD56^dim^) and myeloid DCs in the blood of natalizumab-treated patients, while population of plDCs was proportionally decreased. The stronger inhibition of migration of B cells as compared to monocytes resulted in strong increase in B cell/monocyte ratio in the blood of natalizumab-treated subjects.

**Figure 8:**
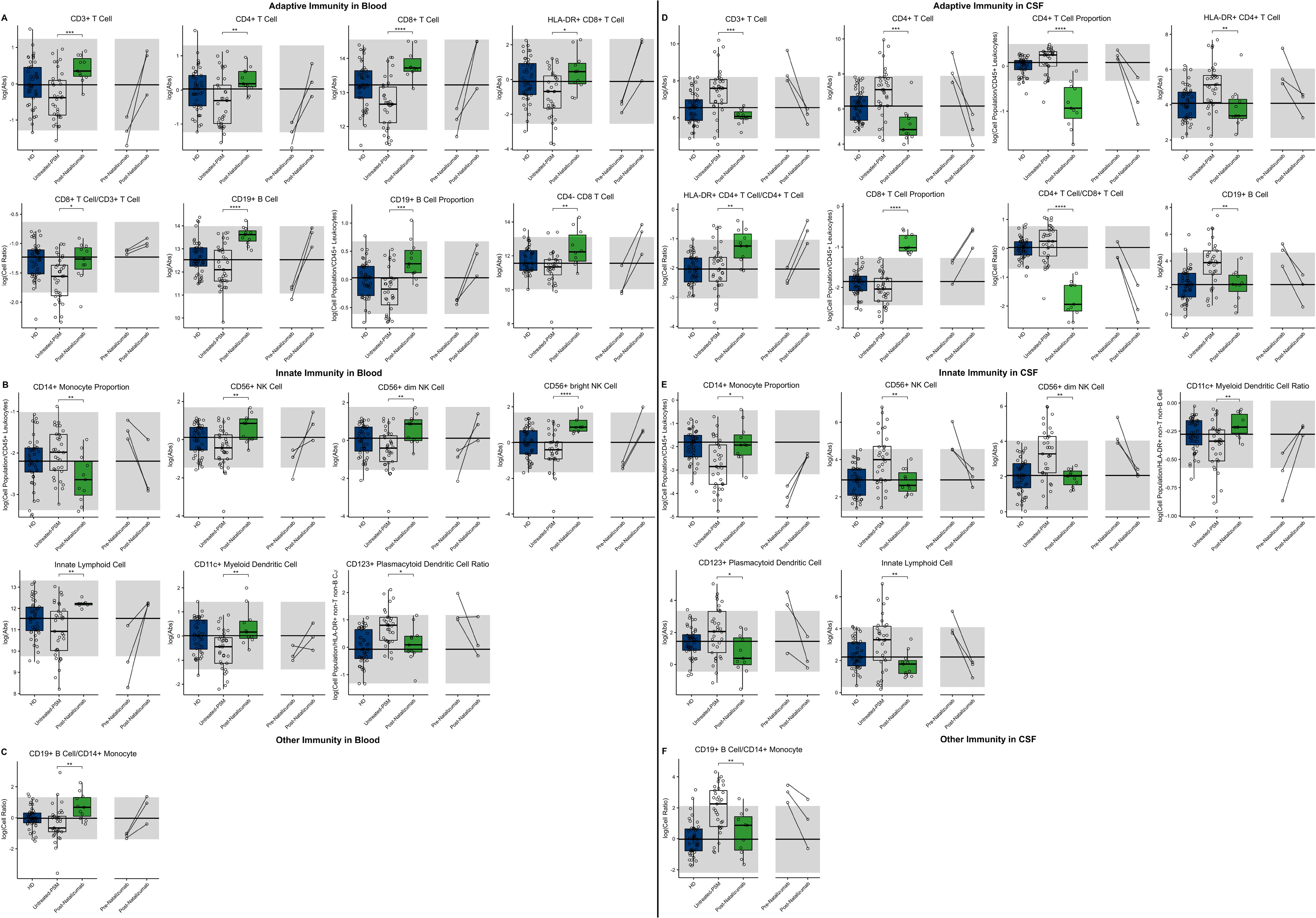
Unpaired t-test with adjustment for multiple comparisons was performed for features during comparison of 33 propensity score matched (PSM) untreated MS patients and 11 natalizumab treated MS patients in blood and CSF. All significant markers were graphed and supplemented with HD cohort and data from 3 longitudinal patients with paired pre-treatment and post-treatment data. \**p* < 0.05, **0.01 *<p* < 0.005, \*\*\**p* < 0.001, \*\*\*\**p* < 0.0001. HD median for each feature was also graphed horizontally and grey shading added representing ± 2 SDs of each feature in HD cohort. (A) Features in adaptive immunity in blood. (B) Features in innate immunity in blood. (C) Features in other immunity in blood. (D) Features in adaptive immunity in CSF. (E) Features in innate immunity in CSF. (F) Features in other immunity in CSF.

Reciprocally, all of the afore-mentioned populations of immune cells were severely depleted from the CSF of natalizumab-treated MS patients, usually significantly below physiological levels. We reproduced stronger inhibitory activity of natalizumab on migration of CD4+ T cells as opposed to CD8+ T cells, resulting in highly non-physiological proportional enrichment of CD8+ T cells over CD4+ T cells in the CSF. On the other hand, the levels of remaining immune cells (i.e., B cells, monocytes, NK cells and DCs) were normalized in the CSF by natalizumab. Only absolute numbers of ILCs fell below physiological levels upon natalizumab treatment.

Again, examination of CSF/blood ratios (Supplementary Figure 4) only highlighted natalizumab’s MOA of inhibiting migration of immune cells from the blood to CSF. The hierarchy of inhibition confirmed much stronger effect on CD4+ as compared to CD8+ T cells, but also highlighted strong effect on all cells of the innate immunity.

#### 3.3.4 Daclizumab

In the blood (Figure 9; Supplementary Presentation 1, Slide 9), daclizumab mildly decreased the absolute numbers and proportions of all lymphocytes of adaptive immunity, while significantly expanding CD56^bright^ NK cells and decreasing numbers of ILCs.

**Figure 9:**
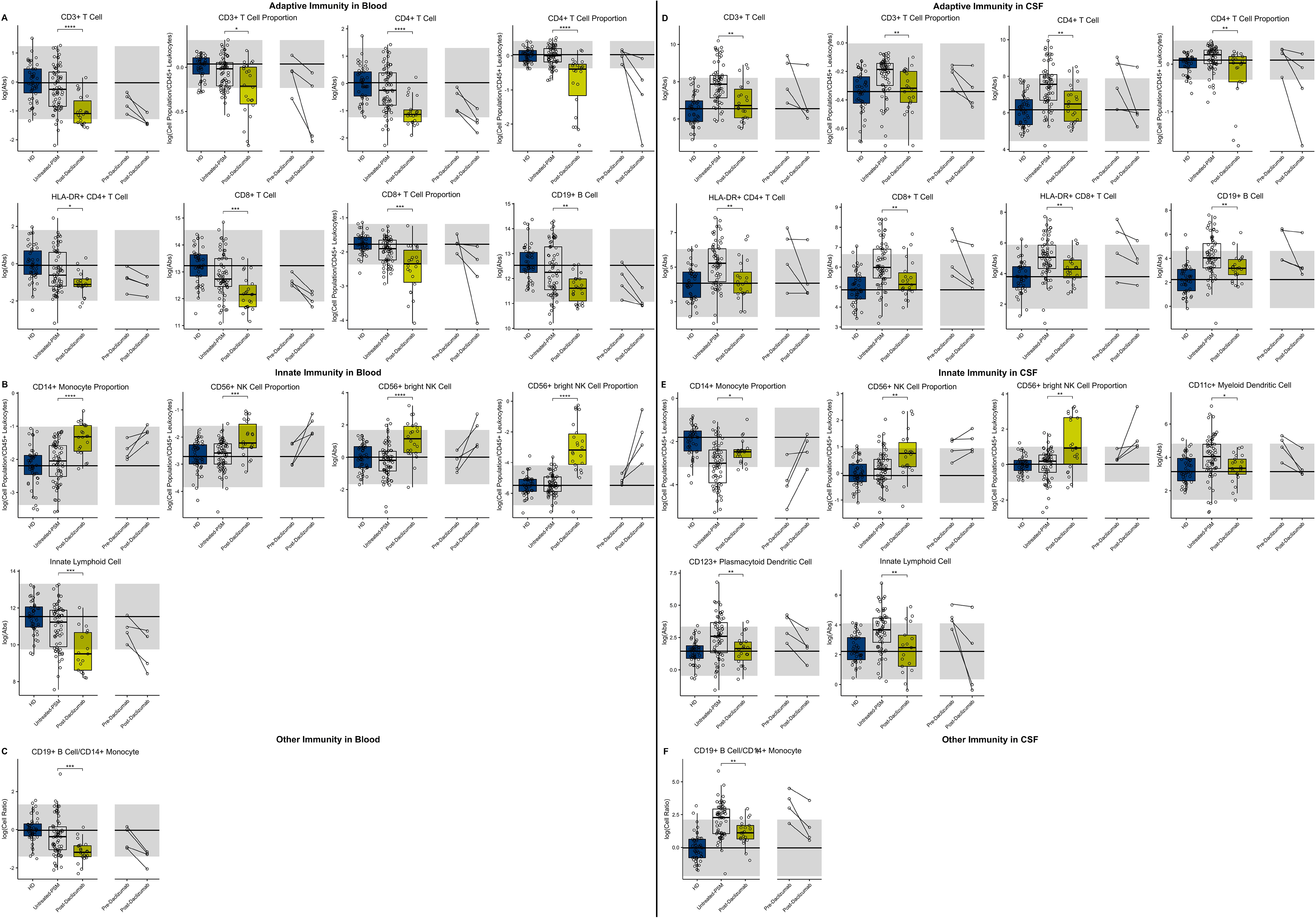
Unpaired t-test with adjustment for multiple comparisons was performed for features during comparison of 66 propensity score matched (PSM) untreated MS patients and 22 daclizumab treated MS patients in blood and CSF. All significant markers were graphed and supplemented with HD cohort and data from 4 longitudinal patients with paired pre-treatment and post-treatment data. \**p* < 0.05, **0.01 *<p* < 0.005, *\*\*\*p* < 0.001, *\*\*\*\*p* < 0.0001. HD median for each feature was also graphed horizontally and grey shading added representing ± 2 SDs of each feature in HD cohort. (A) Features in adaptive immunity in blood. (B) Features in innate immunity in blood. (C) Features in other immunity in blood. (D) Features in adaptive immunity in CSF. (E) Features in innate immunity in CSF. (F) Features in other immunity in CSF.

In the CSF, daclizumab normalized all MS-associated abnormalities except for proportional expansion of CD56^bright^ NK cells that increased above physiological levels seen in HD. Daclizumab also normalized CSF levels of both DC subsets, while proportion of monocytes was increased by daclizumab, but still did not reach HD levels. Because daclizumab had stronger inhibitory effect on CSF T cells compared to CSF B cells and failed to completely normalize monocyte numbers, the CSF B cell/monocyte ratio was significantly decreased by daclizumab therapy but remained above HD levels.

CSF/blood ratios (Supplementary Figure 5) showed that similarly to what was observed for IFN-beta, CD56^bright^ NK cell expansion by daclizumab was much higher in the blood than in the CSF, so that CSF/blood ratio of CD56^bright^ NK cells actually decreased on daclizumab therapy. Finally, we observed that absolute numbers of CSF CD8+ T cells and B cells, as well as B cell/monocyte ratio showed statistically significant negative correlation with length of treatment, suggesting that the efficacy of daclizumab on these markers of CSF inflammation associated with MS increased with the length of therapy.

#### 3.3.5 Ocrelizumab

As expected, ocrelizumab treatment dramatically depleted blood B cells significantly below physiological levels (Figure 10; Supplementary Presentation 1, Slide 10). But Ocrelizumab also exerted milder, nevertheless highly significant effects on other immune cell populations in the blood: increase absolute numbers and/or proportions of most T cell populations and most cells of innate immunity in comparison to propensity score matched untreated MS subjects. While most of the paired before-after blood samples demonstrated same trend, the changes were not congruent in all subjects.

In the CSF, ocrelizumab depleted B cells (again below physiological levels) and normalized most other MS-associated CSF abnormalities. Surprisingly, the inhibitory effect of ocrelizumab was much stronger on CD4+ as compared to CD8+ T cells, so that the resulting proportion of CD8+ T cells rose above physiological levels and CD4/CD8 ratio declined below HD values. MS- associated CSF abnormalities of the innate immunity were normalized by ocrelizumab therapy, including proportion of monocytes.

**Figure 10:**
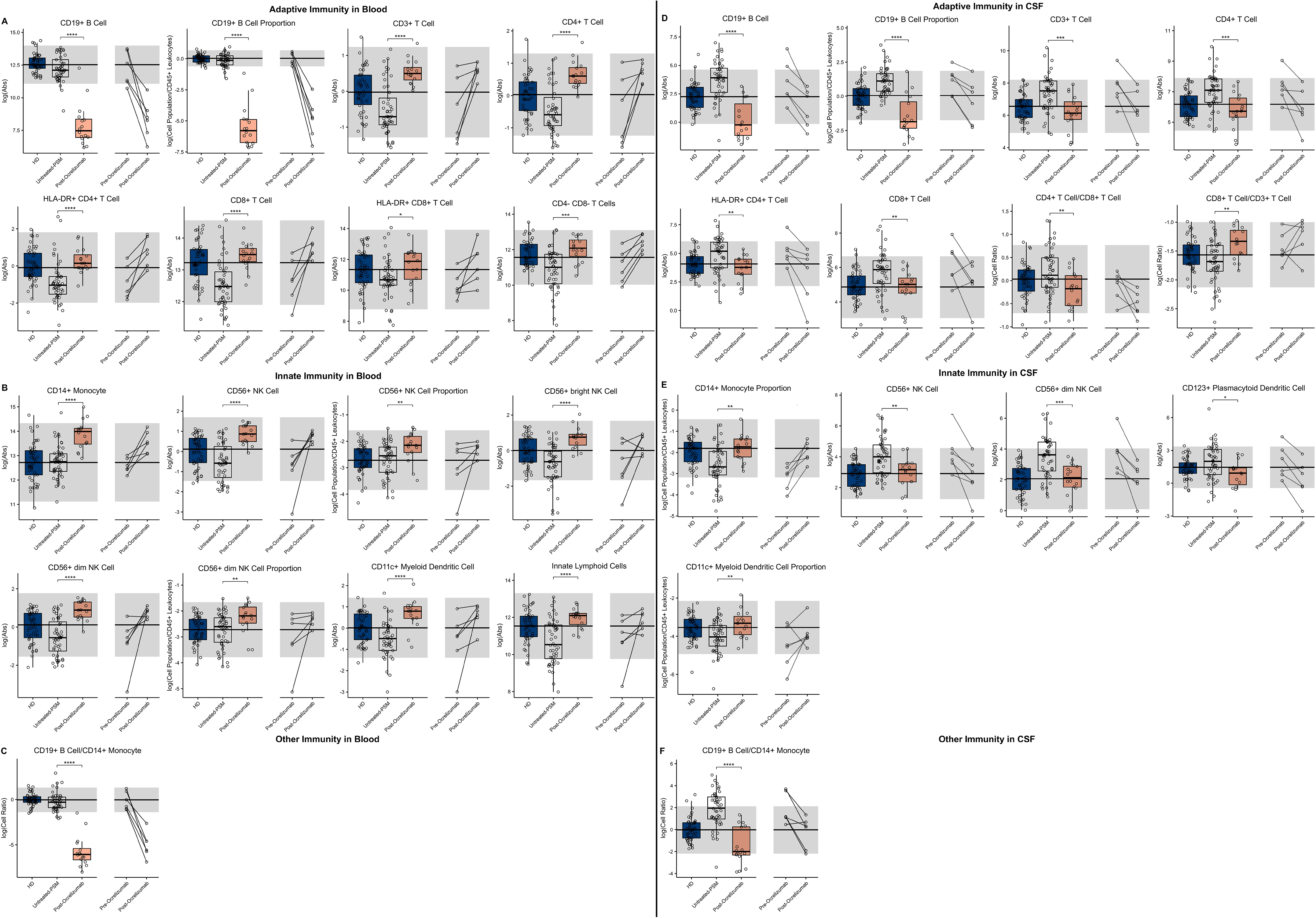
Unpaired t-test with adjustment for multiple comparisons was performed for features during comparison of 48 propensity score matched (PSM) untreated MS patients and 16 ocrelizumab treated MS patients in blood and CSF. All significant markers were graphed and supplemented with HD cohort and data from 6 longitudinal patients with paired pre-treatment and post-treatment data. \**p* < 0.05, **0.01 *<p* < 0.005, \*\*\**p* < 0.001, \*\*\*\**p* < 0.0001. HD median for each feature was also graphed horizontally and grey shading added representing ± 2 SDs of each feature in HD cohort. (A) Features in adaptive immunity in blood. (B) Features in innate immunity in blood. (C) Features in other immunity in blood. (D) Features in adaptive immunity in CSF. (E) Features in innate immunity in CSF. (F) Features in other immunity in CSF.

Perhaps most importantly, out of all drugs studied, ocrelizumab exerted most consistent and broadest treatment duration effects (Figure 11): the absolute number of recently activated HLA-DR+ CD4+ T cells, as well as B cells decreased significantly with the treatment duration, while proportion of CSF monocytes, normally strongly decreased in untreated MS increased in the CSF of ocrelizumab-treated patients, positively correlating with treatment duration.

**Figure 11:**
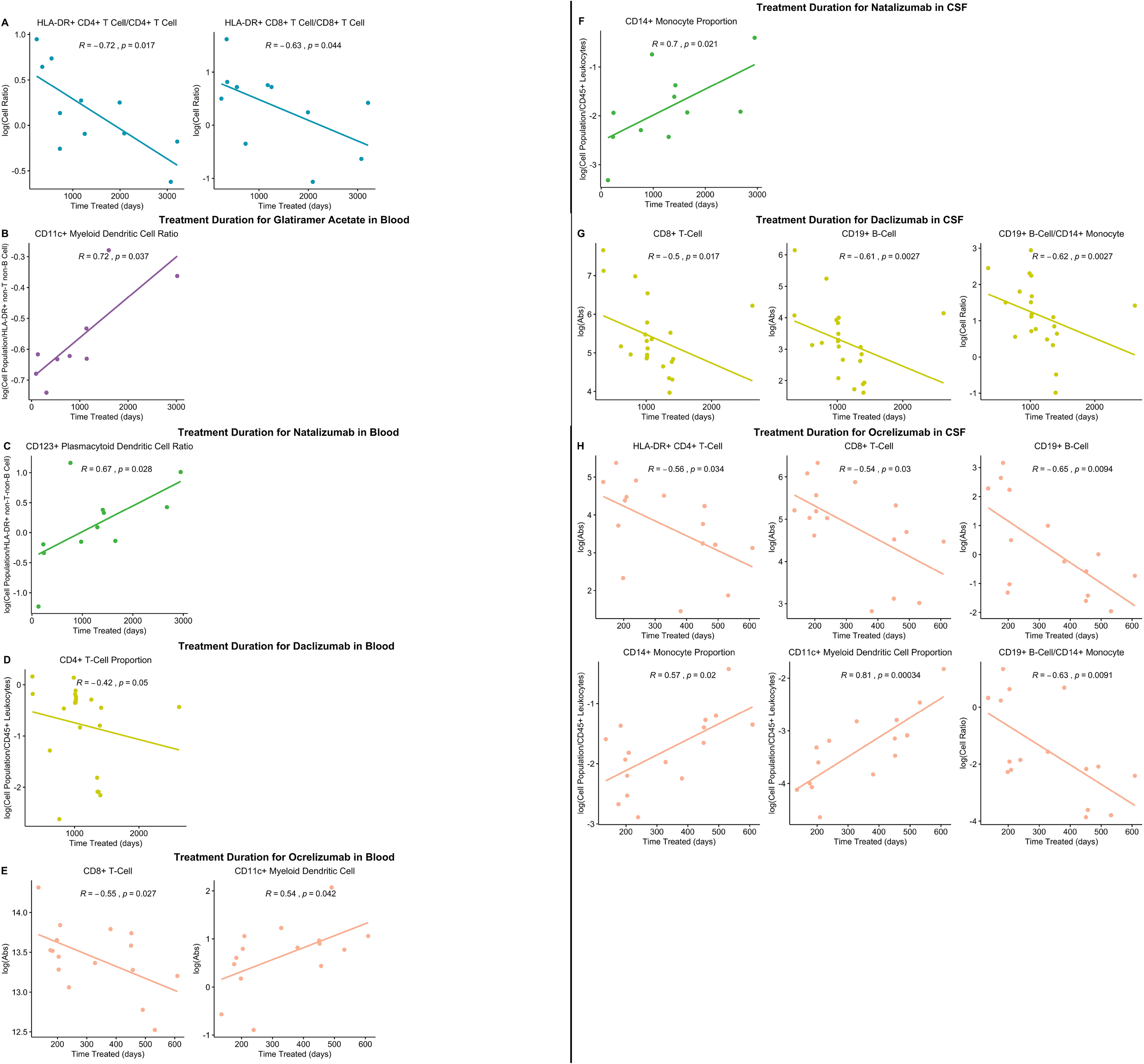
All significantly different features between propensity score matched untreated and treated patients were correlated with treatment duration in treated patients and significant correlations graphed (*p*-value ≤ 0.05). (A) Significant correlations in interferon-beta treated patient blood. (B) Significant correlations in glatiramer acetate treated patient blood. (C) Significant correlations in natalizumab treated patient blood. (D) Significant correlations in daclizumab treated patient blood. (E) Significant correlations in ocrelizumab treated patient blood. (F) Significant correlations in natalizumab treated patient CSF. (G) Significant correlations in daclizumab treated patients CSF. (H) Significant correlations ocrelizumab treated patient in CSF.

Finally, examining CSF/blood ratios (Supplementary Figure 6) showed much stronger B cell depletion in the blood as compared to CSF, while remaining ocrelizumab-induced changes were more prominent in the CSF, arguing that ocrelizumab may be targeting the essential MS-linked biology.

## 4 Discussion

We would like to start by acknowledging the limitations of current study: 1. Due to prospective, longitudinal (i.e., spanning >6 years) design, we could not employ recently developed automated flow cytometry analysis techniques (21). Automated algorithms require identical assay parameters, practically achieved by performing only a single, or highly limited assay run(s) on cryopreserved samples. This is problematic for CSF applications, as small number of CSF cells is poorly amenable to cryopreservation and cryopreservation affects numbers/phenotype of activated immune cells in unpredictable ways. Obvious advantage of the unsupervised clustering algorithms, limitation of bias, was achieved in this study by use of SOP and strict blinding of the personnel involved in the biological samples processing and running the assay. The data were analyzed weekly and after quality control, deposited to research database and locked to prevent further modification. Another advantage of automated clustering is machine learning-aided identification of novel, usually rare cellular populations, possibly related to studied process. We analyzed SOP pre-defined, rare subpopulations of immune cells (e.g., c-kit+ DCs and ILCs, CD56+ monocytes) using isotype-guided gating strategy but failed to validate reproducible differences in these rare subpopulations between diagnostic categories. We believe that key limitation for this effort is small numbers of available CSF cells, even when we process >20cc of CSF and perform immunophenotyping immediately. Because this scarcity of cells would be even more pronounced for cryopreservation-based projects, we consider this limitation unsurmountable. Finally, automated methods provide different results based on the algorithm employed (22). Thus, for CSF applications, both automated and manual/blinded approaches have pros and cons and should be viewed as complimentary. 2. Our results are based on natural history protocol, where only first LP (usually in the untreated stage at the time of diagnostic work-up) was a required procedure and the remaining LPs were optional. This led to an uneven representation of treatments captured in this study and limited number of paired, before /after CSF samples. To mitigate this limitation, we used propensity score matching for formal statistical analyses and provide paired before/after treatment results to assess congruency with group comparisons. The fact that this methodology reproduced vast majority of published observations of drug-related changes from other investigators (4,23-34) support validity of our approach. 3. While we used the independent-validation cohort to validate immunophenotyping features that differed between MS patients and controls, we had inadequate quantities of biological samples to use the same strategy for HD age/gender effects and to validate treatment-induced changes. Thus, novel findings from these unvalidated experiments require independent validation before they can be considered definite, even though all our *p*-values are adjusted for multiple comparisons and we observed good congruency between propensity scored matched group analyses and paired before-after therapy samples. 4. Some of the analyzed cell subpopulations were enumerated based on “exclusion” gates: e.g., CD4-/CD8- CD3+ T cells, or innate lymphoid cells (ILCs). We attempted to identify potentially MS-relevant subpopulations of these “negatively-gated” cells using positive markers such as γδ T cell receptor (TCR) on double-negative T cells and c-kit (FLT3) on ILCs. However, we observed statistically significant differences only in the “parent”, negatively gated cells, but not in the positively gated subpopulations. Again, we believe that this is caused by the scarcity of these minute cell subpopulations in the CSF, even under moderate inflammation associated with MS. Although negatively gated parent cells had to express ubiquitous marker of immune cells CD45 and be localized within lymphocyte or monocyte forward/side scatter gate, lack of definite positive identification marker of these cells makes us extra cautious in our interpretations. Therefore, we decided to omit these results in the summary models and associated video, even though we did include them in all other Figures and results.

Notwithstanding these limitations, our results broaden understanding of the evolution of immune cellular changes in blood and CSF compartments across HD and MS lifespans and provide novel, comparative analyses of several MS treatment modalities.

While we reproduced previously published differences between blood and CSF immune cellular compositions (3,35-41), to our best knowledge, the physiological effects of gender and age on CSF cellular composition have not been previously defined and represent first novel aspects of this study. Despite the lack of independent validation, our ability to reproduce previously published healthy age/gender effects on *blood* immune cells within identical HD cohort strengthens the probability that *CSF* data will be, likewise, reproducible in future studies. These reproduced age changes in blood include declines in proportion of plDCs with age (42), increases in CD56+ NK cells (43-46) with age, positive correlation of HLA-DR+ CD8+ T cells (47,48) and HLA-DR+ CD4+ T cells (47) with age, increases in CD4+ T cell/CD3+ T cell ratio with age (49), and increases in proportion of CD3+ T cells (50,51) and proportion of CD4+ T cells with age (51). Reproduced gender differences in blood include higher presence of B cells in females (52), higher CD4/CD8 ratio in females (53,54), higher proportion of CD8+ T cells in males (54), and higher presence of HLA-DR+ CD4+ T cells and HLA-DR+ CD8+ T cells in males (55).

The physiological gender/age effects on cellular composition of the *CSF* were more limited: we observed higher CD4/CD8 ratio and greater proportion of NK cells in female as compared to male HDs. Physiological aging was associated with lower absolute counts of CSF B cells, higher proportion of CD4+ T cells and, quite surprisingly, lower blood/CSF ratios of HLA-DR+ T cells, both CD4+ and CD8+. This latter change is surprising in view of the higher *blood* levels of activated, HLA-DR+ T cells associated with aging ((48); validated only for HLA-DR+ CD8+ T cells here) and implies decreased migration of these recently activated T cells from blood to CSF. Thus, we conclude that there is likely age-related decrease in immunosurveillance of CNS tissue, which may, at least partially underlie the epidemiological observations of second peak of CNS malignancies (56,57) and high morbidity of CNS infections in aged population (58,59).

After adjusting for physiological gender and age effects and analyzing only samples from untreated subjects, we identified (and validated in the independent cohort) MS-specific changes in the blood: increased proportion of CD4+ T cells (among all T cells) observed in all MS subtypes and associated increase in CD4/CD8 ratio, even though the latter validated statistical significance only in the PMS cohort. Several previous studies found either higher CD4+ T cell counts/proportions in untreated MS (35,36,60) or increased proportions of *activated* CD4+ T cells in MS (61-63), using variety of different activation markers. Although our study cannot address causal relationships, the fact that we observed statistically significant increase in proportion of CD4+ T cells and CD4/CD8 ratio with disease duration in our untreated MS cohort suggest that these changes may be a consequence of MS disease process. Indeed, a reciprocal association of declining proportion of CD4+ T cells with MS DD was observed in the CSF. There are several interpretations for this reciprocity: 1. Decreased recruitment of CD4+ T cells from blood to CSF or, 2. Increased recruitment of CD8+ T cells from blood to CSF. Additionally, we must remember that immune cells in the CSF do not equate cellular composition of CNS tissue once the compartmentalized inflammation was established (64). Indeed, the CSF concentrations of immune cell-specific cell surface markers, such as CD27 (predominantly shed from T cells especially CD8+), CD21 (shed from B cells, especially naive B cells), CD23 (predominantly shed by activated DCs and memory B cells) and CD14/CD163 (predominantly shed by monocytes and macrophages) are equally increased in all MS subtypes (64), indicating similar levels of inflammation in the intrathecal compartment of RRMS and PMS subjects. In fact, the ratio between these shed cell-surface markers and the numbers of the cells of their origin in the CSF represents *in-vivo* measure of inflammation compartmentalized to CNS tissue, which demonstrated significantly higher CNS compartmentalization of inflammation in PMS in comparison to RRMS (64). This in vivo observation reproduces MS pathology studies (65,66).

Consequently, a unifying explanation for most observations in the current study is increased recruitment and enhanced retention of immune cells in CNS tissue (i.e., compartmentalized inflammation) as MS evolves. Additional results supporting this unified explanation are: 1. In PMS blood, we identified decrease in absolute numbers and proportions of T cells, affecting all T cell subtypes, including recently activated, HLA-DR+ T cells. This is unexpected, as proportion of these cells in the blood increases with physiological aging ((48) and this study) and due to high prevalence of urinary tract infections in PMS patients one would expect further increase of HLA- DR+ T cells in PMS blood. Because such recently activated effector T cells easily cross blood brain barrier, we consider their recruitment to and retention in CNS likely. In agreement, the CSF/blood ratios of HLA-DR+ T cells is significantly increased in both MS subtypes, but also in OIND, and for HLA-DR+/CD4+ T cells, weakly but significantly, also in NIND controls. This supports the notion that these recently activated T cells are easily recruited to tissues in response to tissue damage. 2. MS patients have increased levels of all measured immune cell subpopulations in the CSF, except for monocytes and granulocytes. This inflammatory reaction is not MS specific; subjects with OIND have even higher concentrations of immune cells in the CSF. However, the new finding is that we see statistically significant decreases of all of these CSF cells with MS duration, not as a dichotomized process separating RRMS from PMS (in fact, no immunophenotyping feature reliably differentiated MS subtypes), but as a continuous process. This data strongly supports MS-driven changes that lead to progressive compartmentalization of the inflammatory process to CNS tissue, such as formation of tertiary lymphoid follicles.

Are these cellular changes, identified and validated in untreated MS population normalized by FDA-approved treatments? Based on our inclusion criteria, we had data to answer this question for 5 drugs. Because of retraction of daclizumab from the markets and previous extensive analyses of the effects of daclizumab on blood and CSF immune cells (25,31), we used daclizumab only for comparisons and will focus the discussion on remaining four drugs: two low efficacy drugs classes (i.e., IFN-beta and glatiramer acetate [GA] preparations) and two high efficacy drugs (i.e., natalizumab and ocrelizumab).

All studied drugs decreased levels of adaptive immunity in the CSF, but they did it to different extent: low efficacy drugs normalized absolute numbers of CSF T cells and CSF B cells, while high efficacy drugs decreased CSF immune levels below physiological levels: natalizumab decreased absolute levels and proportions of CD4+ but not CD8+ T cells below HD levels, while normalizing CSF B cells, monocytes and NK cells. The relative selectivity of natalizumab’s action to CD4+ T cells led to non-physiological decrease in CSF CD4/CD8 ratio and highly non-physiological increase in the proportion of CSF CD8+ T cells. Ocrelizumab predictably decreased CSF B cells below physiological levels and normalized remaining CSF immunophenotyping parameters except for CD4/CD8 T cells ratio, which decreased below physiological levels due to relative increase in proportion of CD8+ T cells. Thus, studied low efficacy drugs normalized MS-associated changes in CSF lymphocytes, whereas high efficacy treatments introduced non-physiological changes with more pronounced decrease in CD4+ T cells as compared to CD8+ T cells and non-physiological decreases in CSF CD4/CD8 ratio. This was not observed with daclizumab, which decreased CSF levels of CD4+ and CD8+ T cells to a similar extent.

The other important differences between drug effects on CSF cellular populations resided in their effects on HLA-DR+ T cells: in general, low efficacy drugs failed to inhibit these recently activated T cells. In fact, IFN-beta significantly increased proportion of both CD4+ and CD8+/HLA-DR+ T cells and GA increased proportion of CD4+/HLA-DR+T cells. The effect of IFN-beta on these cells is unique, as it also increased both absolute numbers and proportions of HLA-DR+ CD4+ and CD8+ T cells in the blood, arguing for drug-specific effect that may not be beneficial for MS. Actually, our human observations agree with anti-apoptotic effect on activated T cells of type-1 interferons described in animals (23). This suggest that IFN-beta may have both beneficial effects for MS, such as stabilization of blood-brain barrier and increasing proportions of regulatory CD56^bright^ NK cells (30,32), while its unwanted effects include persistence of activated T cells, which may enhance anti-viral immunity, but may be detrimental in MS.

Natalizumab also failed to normalize HLA-DR+ T cells: the proportion of CD4+/HLA-DR+ T cells increased in the CSF of natalizumab-treated patients, either due to lower inhibition of their migration from blood, or due to their subsequent expansion in the lymphocyte-depleted intrathecal compartment. Only ocrelizumab (and daclizumab) normalized CSF numbers of HLA-DR+ T cells. Perhaps most importantly, we observed significant correlations between reversal of CSF immunophenotyping abnormalities and duration of treatment for ocrelizumab (and, to a lesser degree, for daclizumab). Other tested drugs did not exhibit significant relationships between treatment duration and normalization of MS-related immunophenotyping abnormalities. This suggests that efficacy of ocrelizumab increases with treatment duration, at least for 2 years, which was the longest CSF follow-up in our cohort. This result is important for the long-term use of ocrelizumab in MS.

When it comes to cells of innate immunity, low efficacy drugs normalized CSF levels of myDC (IFN-beta only) and plDC (both drugs) but failed to normalize CSF levels of NK cells and monocytes. GA exerted beneficial and statistically significant effect on monocytes, but median levels of CSF monocytes in the treated group were still below HD median. IFN-beta additionally increased levels/proportions of immunoregulatory CD56^bright^ NK cells, both in the blood and CSF, consistent with published studies (30,32). The high efficacy drugs had almost identical effects on CSF innate immunity: both normalized numbers of NK cells in the CSF (especially cytotoxic CD56^dim^ NK cells) and increased proportion of monocytes. They also decreased numbers of CSF plDCs but had no significant effects on myDCs. Thus, three MS drugs exerted higher inhibitory effect on plasmacytoid as compared to myeloid DCs (i.e., GA, natalizumab and ocrelizumab), whereas IFN-beta preparations and daclizumab inhibited both DC subsets equally. Interestingly, these two drugs also significantly expanded CD56^bright^ NK cells, both in the blood and CSF, even though daclizumab exerted stronger effects. It would be interesting to explore functional relationships between CD56^bright^ NK cells and myDCs in future MS studies.

The analysis of the drug’s effects on cell populations in the blood and on blood/CSF ratios provide additional insight into MOA of these drugs. GA increased blood levels of CD8+ T cells, CD4- /CD8- T cells and CD56^bright^ NK cells. All these cell types were decreased in untreated MS, especially PMS, in comparison to HDs. GA also normalized CSF/blood ratios for all cells of adaptive immunity. Thus, we interpret these changes as normalization of MS-associated disturbances in blood immune cells. In contrast, IFN-beta exerted several non-physiological blood changes. In addition to previously mentioned increases in HLA-DR+ CD4+ and CD8+ T cells and CD56^bright^ NK cells, IFN-beta preparations also increased number of blood B cells above physiological levels and significantly decreased proportion of blood myDCs. These changes are consistent with known direct and indirect activating effects of type-1 interferons on immune cells: e.g., the stimulation of proliferation of NK cells and T cells is IL-15 driven (34), explaining preferential expansion of CD56^bright^ NK cells and HLA-DR+ T cells that express high levels of CD122 and CD132, which together constitute intermediate-affinity IL-2 receptor, used for IL-15 trans-presentation.

By inhibiting migration of immune cells from blood to tissues, natalizumab induces non-physiological increases of blood numbers of most cells of adaptive immunity and both types of NK cells, even though the effect on CD56^bright^ NK cells is more robust in comparison to CD56^dim^ NK cells. In contrast, migration of cells of myeloid lineage, such as monocytes and granulocytes, is not significantly inhibited by natalizumab and their blood numbers are not affected. The highly non-physiological effects of natalizumab on CSF/blood ratios, not shared by remaining drugs studied in this paper, reflects its known MOA: inhibiting cell migration from blood to tissues, with stronger inhibition of CD4+ T cell in comparison to CD8+ T cells, leading to strong proportional enrichment of CD4+ T cells in the blood and CD8+ T cells in the CSF of treated patients (4). The significant effects on CSF/blood ratios of both types of NK cells, both types of DCs and ILC support inhibitory action of natalizumab on extravasation of these innate immune cells CSF. As expected, ocrelizumab caused selective non-physiological depletion of blood B cells. This was associated with normalization and/or even overshoot of MS-associated disturbances in cellular blood composition, such as increase in absolute numbers of blood T cells, both CD4+ and CD8+ and their HLA-DR+ subpopulations, but also blood monocytes and NK cells (CD56^bright^>CD56^dim^). Because the absolute numbers of most of these cells increased in the blood and simultaneously decreased in the CSF, CSF/blood ratios mostly decreased below physiological levels. The striking exceptions were CSF/blood ratios of B cells. B cells were depleted in both compartments, but proportionally more in the blood, causing paradoxical increase in CSF/blood B cell ratio above the untreated matched MS patients.

In conclusion, our results support the notion that MS is a single disease, with clinical categories of RRMS, SPMS and PPMS representing different stages of a continuous disease evolution. From the standpoint of cellular immunity, the MS evolution is characterized by constant recruitment of activated T cells to CNS tissue and establishment of environment conducive to compartmentalized inflammation. All studied MS drugs inhibit cellular inflammatory CSF responses in comparison to untreated MS. From all studied drugs, GA induces least non-physiological changes in cellular immunity, while achieving comparable, if not better inhibition of CSF inflammation in comparison to IFN-beta preparations. IFN-beta have a disadvantage of inducing chronic activation of the immune system. High efficacy drugs exert stronger anti-inflammatory effects in CSF but induce many non-physiological changes that may underlie associated risk of infectious complications or cancers under long-term use. Natalizumab is clearly worse in this regard than ocrelizumab. The efficacy of ocrelizumab on CSF inflammatory response increases with the length of treatment, at least for 2 years. The difference between high and low efficacy drugs on disability progression likely reflects more potent inhibition of inflammation in CNS tissue, even though this could not be addressed in the current study.

The associated video that integrates major results of this study may be used for education.

## Data Availability

The datasets presented in this study can be found in online repositories.

https://github.com/vanessatmorgan/Bielekova-Lab-Code/tree/master/FormerLabMembers/Paav

## 5 Acknowledgements

We thank all the Neuroimmunological Diseases Section (NDS) postbaccalaureate research fellows who performed flow cytometry, including - but not limited to - Ann Marie Weideman, Alexandra Boukhvalova, Kayla Jackson, Linh Pham, and Jon Phillips. From the wet lab, we also thank Elena Romm for processing samples and Ruturaj Masvekar for helping with manuscript preparation. From the clinical side, we also thank our clinicians - Alison Wichman and Mary Sanford - for performing lumbar punctures and seeing patients at the clinic, our patient care coordinator Michelle Woodland, and our regulatory nurse Tiffany Houser. Finally, we thank our patients and their caregivers for partnering with us on this research.

## 6 Author Contributions Statement

BB designed the study and guided/supervised all its aspects; PH contributed to data collection and analysed the data; PK contributed to quality control and maintenance of data and contributed to data analysis; CB exported data and contributed to data analysis; BB and PH wrote the manuscript; all authors contributed to manuscript revision, read and approved the submitted version.

## 9 Data Availability

The datasets presented in this study can be found in online repositories. The names of the repository/repositories and accession number(s) can be found below: https://github.com/vanessatmorgan/Bielekova-Lab-Code/tree/master/FormerLabMembers/Paav.

## 13 Supplementary Materials

**Supplementary Presentation 1:** Major results of this study are described in this presentation, beginning with description of immune cell populations in healthy blood and CSF, and further narration of age and gender effects on these populations in HDs. Then RRMS and PMS patient blood and CSF immune system physiologies are differentiated from HDs. Lastly, interferon-beta, glatiramer acetate, natalizumab, daclizumab, and ocrelizumab treated MS patients are compared to untreated in MS patients in this 15-minute video.

**Supplementary Video 1:** Major results of this study are described in this educational video beginning with description of immune cell populations in healthy blood and CSF, and further narration of age and gender effects on these populations in HDs. Then RRMS and PMS patient blood and CSF immune system physiologies are differentiated from HDs. Lastly, interferon-beta, glatiramer acetate, natalizumab, daclizumab, and ocrelizumab treated MS patients are compared to untreated in MS patients in this 15-minute video.

**Supplementary Figure 1:** Features that validated in the independent validation cohort in CSF and/or blood were calculated as CSF/blood ratios. CSF/blood ratios were then compared between HDs and RRMS and PMS patients, and statistically significant features graphed. HDs, NINDs, OINDs, RRMS, and PMS cohorts for each feature were graphed. Unpaired t-test was completed to compare each cohort to HDs with adjustment for multiple comparisons. \**p* < 0.05, **0.01 *<p* < 0.005, \*\*\**p* < 0.001, \*\*\*\**p* < 0.0001. HD median for each feature was also graphed horizontally and grey shading added representing ± 2 SDs of each feature in HD cohort. *A*. Features in adaptive immunity. *B*. Features in innate immunity. *C*. Features in other immunity. *D*. Features that validated were correlated with disease duration and statistically significant features graphed (*p*- value ≤ 0.05). MS subtypes were color coded to show heterogeneity of cohorts in each significant feature, with blue representing RRMS, red representing PPMS, and orange representing SPMS patients.

**Supplementary Figure 2:** Features that were statistically significant in blood and/or CSF between propensity score matched (PSM) untreated and interferon-beta treated MS patients were calculated as CSF/blood ratios. CSF/blood ratios were then compared between 36 propensity score matched untreated and 12 interferon-beta treated MS patients and an unpaired t-test with adjustment for multiple comparisons performed. All significant markers were graphed and supplemented with HD cohort and data from 4 longitudinal patients with paired pre-treatment and post-treatment data. \**p* < 0.05, **0.01 *<p* < 0.005, \*\*\**p* < 0.001, \*\*\*\**p* < 0.0001. HD median for each feature was also graphed horizontally and grey shading added representing ± 2 SDs of each feature in HD cohort. *A*. Features in adaptive immunity. *B*. Features in innate immunity.

**Supplementary Figure 3:** Features that were statistically significant in blood and/or CSF between propensity score matched (PSM) untreated and glatiramer acetate treated MS patients were calculated as CSF/blood ratios. CSF/blood ratios were then compared between 33 propensity score matched untreated and 11 glatiramer acetate (GA) treated MS patients and an unpaired t-test with adjustment for multiple comparisons performed. All significant markers were graphed and supplemented with HD cohort and data from 5 longitudinal patients with paired pre-treatment and post-treatment data. \**p* < 0.05, **0.01 *<p* < 0.005, \*\*\**p* < 0.001, \*\*\*\**p* < 0.0001. HD median for each feature was also graphed horizontally and grey shading added representing ± 2 SDs of each feature in HD cohort. *A*. Features in adaptive immunity. *B*. Features in innate immunity. *C*. Features in other immunity.

**Supplementary Figure 4:** Features that were statistically significant in blood and/or CSF between propensity score matched (PSM) untreated and natalizumab treated MS patients were calculated as CSF/blood ratios. CSF/blood ratios were then compared between 33 propensity score matched untreated and 11 natalizumab treated MS patients and an unpaired t-test with adjustment for multiple comparisons performed. All significant markers were graphed and supplemented with HD cohort and data from 3 longitudinal patients with paired pre-treatment and post-treatment data. \**p* < 0.05, **0.01 *<p* < 0.005, \*\*\**p* < 0.001, \*\*\*\**p* < 0.0001. HD median for each feature was also graphed horizontally and grey shading added representing ± 2 SDs of each feature in HD cohort. *A*. Features in adaptive immunity. *B*. Features in innate immunity. *C*. Features in other immunity.

**Supplementary Figure 5:** Features that were statistically significant in blood and/or CSF between propensity score matched (PSM) untreated and daclizumab treated MS patients were calculated as CSF/blood ratios. CSF/blood ratios were then compared between 66 propensity score matched untreated and 22 daclizumab treated MS patients and an unpaired t-test with adjustment for multiple comparisons performed. All significant markers were graphed and supplemented with HD cohort and data from 4 longitudinal patients with paired pre-treatment and post-treatment data. \**p* < 0.05, **0.01 *<p* < 0.005, \*\*\**p* < 0.001, \*\*\*\**p* < 0.0001. HD median for each feature was also graphed horizontally and grey shading added representing ± 2 SDs of each feature in HD cohort. *A*. Features in innate immunity.

**Supplementary Figure 6:** Features that were statistically significant in blood and/or CSF between propensity score matched (PSM) untreated and ocrelizumab treated MS patients were calculated as CSF/blood ratios. CSF/blood ratios were then compared between 48 propensity score matched untreated and 16 ocrelizumab treated MS patients and an unpaired t-test with adjustment for multiple comparisons performed. All significant markers were graphed and supplemented with HD cohort and data from 6 longitudinal patients with paired pre-treatment and post-treatment data. \**p* < 0.05, **0.01 *<p* < 0.005, \*\*\**p* < 0.001, \*\*\*\**p* < 0.0001. HD median for each feature was also graphed horizontally and grey shading added representing ± 2 SDs of each feature in HD cohort. *A*. Features in adaptive immunity. *B*. Features in innate immunity. *C*. Features in other immunity.

**Supplementary Table 1:** Antibodies used in flow cytometry are described here with targets, dyes, clones, manufacturers, and product numbers.

| Target: | Dye: | Clone: | Manufacturer: | Product Number: |
| --- | --- | --- | --- | --- |
| CD3 | Qdot 655 | s4.1 | Invitrogen by Thermo Scientific | Q10012 |
| CD4 | Qdot 705 | s3.5 | Invitrogen by Thermo Scientific | Q10060 |
| CD8 | AmCyan | SK1 | BD | 339188 |
| CD11c | PE-Cyanine7 | 3.9 | eBioscience | 25-0116-42 |
| CD14 | Alexa Fluor 700 | HCD14 | BioLegend | 325614 |
| CD19 | BV605 | SJ25C1 | BD Horizon | 562653 |
| CD45 | V450 | HI30 | BD Horizon | 560367 |
| CD56 | FITC | TULY56 | Invitrogen by Thermo Scientific | 11-0566-42 |
| Ckit | PE | 47233 | R&D Systems | FAB332P |
| CD123 | PerCP-Cyanine5.5 | 6H6 | eBioscience | 45-1239-42 |
| TCR-gd | APC | B1 | BD Pharmingen | 555718 |
| HLA-DR | APC-eFluor 780 | LN3 | eBioscience | 47-9956-42 |

**Supplementary Table 2:** Untreated MS patient cohort was divided into independent training and validation cohorts with a 50/50 split, while accounting for age, gender, and MS subtypes. Demographics of these cohorts can be observed.

|  | Training Cohort: | Validation Cohort: |
| --- | --- | --- |
| **Number of Patients** | 59 | 59 |
| **Age (mean ± SD)** | 49.33 ± 12.88 | 52.14 ± 12.62 |
| **Male/Female** | 24/35 | 24/35 |
| **CombiWISE (mean ± SD)** | 39.59 ± 21.22 | 38.88 ± 18.79 |
| **EDSS (mean ± SD)** | 4.72 ± 2.31 | 4.66 ± 1.98 |
| **RRMS/PMS** | 16/43 | 19/40 |

## References

1. Höftberger R. Neuroimmunology: An expanding frontier in autoimmunity. Front Immunol (2015) doi :10.3389/fimmu.2015.00206

2. Gastaldi M, Zardini E, Scaranzin S, Uccelli A, Andreetta F, Baggi F, Franciotta D. Autoantibody Diagnostics in Neuroimmunology: Experience From the 2018 Italian Neuroimmunology Association External Quality Assessment Program. Front Neurol (2020) doi:10.3389/fneur.2019.01385

3. Han S, Lin YC, Wu T, Salgado AD, Mexhitaj I, Wuest SC, Romm E, Ohayon J, Goldbach-Mansky R, Vanderver A, et al. Comprehensive Immunophenotyping of Cerebrospinal Fluid Cells in Patients with Neuroimmunological Diseases. J Immunol (2014) doi :10.4049/jimmunol.1302884

4. Stüve O, Marra CM, Bar-Or A, Niino M, Cravens PD, Cepok S, Frohman EM, Phillips JT, Arendt G, Jerome KR, et al. Altered CD4+/CD8+ T-cell ratios in cerebrospinal fluid of natalizumab-treated patients with multiple sclerosis. Arch Neurol (2006) doi:10.1001/archneur.63.10.1383

5. Eggers EL, Michel BA, Wu H, Wang SZ, Bevan CJ, Abounasr A, Pierson NS, Bischof A, Kazer M, Leitner E, et al. Clonal relationships of CSF B cells in treatment-naive multiple sclerosis patients. JCI insight (2017) doi:10.1172/jci.insight.92724

6. Pitteri M, Ziccardi S, Dapor C, Guandalini M, Calabrese M. Lost in classification: Lower cognitive functioning in apparently cognitive normal newly diagnosed RRMS patients. Brain Sci (2019) doi:10.3390/brainsci9110321

7. Gökdoğan Edgünlü T, Ünal Y, Karakaş Çelik S, Genç Ö, Emre U, Kutlu G. The effect of FOXO gene family variants and global DNA metylation on RRMS disease. Gene (2020) doi:10.1016/j.gene.2019.144172

8. Geraci F, Ragonese P, Barreca MM, Aliotta E, Mazzola MA, Realmuto S, Vazzoler G, Savettieri G, Sconzo G, Salemi G. Differences in intercellular communication during clinical relapse and gadolinium-enhanced MRI in patients with relapsing remitting multiple sclerosis: A study of the composition of extracellular vesicles in cerebrospinal fluid. Front Cell Neurosci (2018) doi:10.3389/fncel.2018.00418

9. Wurth S, Kuenz B, Bsteh G, Ehling R, Di Pauli F, Hegen H, Auer M, Gredler V, Deisenhammer F, Reindl M, et al. Cerebrospinal fluid B cells and disease progression in multiple sclerosis - A longitudinal prospective study. PLoS One (2017) doi: 10.1371/journal.pone.0182462

10. Monson NL, Cravens PD, Frohman EM, Hawker K, Racke MK. Effect of rituximab on the peripheral blood and cerebrospinal fluid B cells in patients with primary progressive multiple sclerosis. Arch Neurol (2005) doi:10.1001/archneur.62.2.258

11. Zhang Y, McClellan M, Efros L, Shi D, Bielekova B, Tang MT, Vexler V, Sheridan JP. Daclizumab reduces CD25 levels on T cells through monocyte-mediated trogocytosis. Mult Scler (2014) doi:10.1177/1352458513494488

12. Gingele S, Jacobus T, Konen F, Hümmert M, Sühs K-W, Schwenkenbecher P, Ahlbrecht J, Möhn N, Müschen L, Bönig L, et al. Ocrelizumab Depletes CD20+ T Cells in Multiple Sclerosis Patients. Cells (2018) doi:10.3390/cells8010012

13. Kosa P, Barbour C, Wichman A, Sandford M, Greenwood M, Bielekova B. NeurEx: digitalized neurological examination offers a novel high-resolution disability scale. Ann Clin Transl Neurol (2018) doi:10.1002/acn3.640

14. Kurtzke JF. Rating neurologic impairment in multiple sclerosis: An expanded disability status scale (EDSS). Neurology (1983) doi:10.1212/wnl.33.11.1444

15. Kosa P, Ghazali D, Tanigawa M, Barbour C, Cortese I, Kelley W, Snyder B, Ohayon J, Fenton K, Lehky T, et al. Development of a sensitive outcome for economical drug screening for progressive multiple sclerosis treatment. Front Neurol (2016) doi:10.3389/fneur.2016.00131

16. Polman CH, Reingold SC, Banwell B, Clanet M, Cohen JA, Filippi M, Fujihara K, Havrdova E, Hutchinson M, Kappos L, et al. Diagnostic criteria for multiple sclerosis: 2010 Revisions to the McDonald criteria. Ann Neurol (2011) doi:10.1002/ana.22366

17. Thompson AJ, Banwell BL, Barkhof F, Carroll WM, Coetzee T, Comi G, Correale J, Fazekas F, Filippi M, Freedman MS, et al. Diagnosis of multiple sclerosis: 2017 revisions of the McDonald criteria. Lancet Neurol (2018) doi:10.1016/S1474-4422(17)30470-2

18. Weideman AM, Tapia-Maltos MA, Johnson K, Greenwood M, Bielekova B. Meta-analysis of the age-dependent efficacy of multiple sclerosis treatments. Front Neurol (2017) doi:10.3389/fneur.2017.00577

19. R Studio Team. R Studio. RS *ed* http://www.rstudio.com/(2015) doi:http://www.rstudio.com/.

20. Barbour C, Kosa P, Varosanec M, Greenwood M, Bielekova B. Molecular models of multiple sclerosis severity identify heterogeneity of pathogenic mechanisms. *medRxiv* (2020)2020.05.18.20105932. doi:10.1101/2020.05.18.20105932

21. Mair F, Hartmann FJ, Mrdjen D, Tosevski V, Krieg C, Becher B. The end of gating? An introduction to automated analysis of high dimensional cytometry data. Eur J Immunol (2016) doi:10.1002/eji.201545774

22. Aghaeepour N, Finak G, Hoos H, Mosmann TR, Brinkman R, Gottardo R, Scheuermann RH, Dougall D, Khodabakhshi AH, Mah P, et al. Critical assessment of automated flow cytometry data analysis techniques. Nat Methods (2013) doi:10.1038/NMETH.2365

23. Marrack P, Kappler J, Mitchell T. Type I interferons keep activated T cells alive. J Exp Med (1999) doi:10.1084/jem.189.3.521

24. Rommer PS, Dudesek A, Stüve O, Zettl UK. Monoclonal antibodies in treatment of multiple sclerosis. Clin Exp Immunol (2014) doi:10.1111/cei.12197

25. Lin YC, Winokur P, Blake A, Wu T, Romm E, Bielekova B. Daclizumab reverses intrathecal immune cell abnormalities in multiple sclerosis. Ann Clin Transl Neurol (2015) doi:10.1002/acn3.181

26. Warnke C, Stettner M, Lehmensiek V, Dehmel T, Mausberg AK, Von Geldern G, Gold R, Kümpfel T, Hohlfeld R, Mäurer M, et al. Natalizumab exerts a suppressive effect on surrogates of B cell function in blood and CSF. Mult Scler (2015) doi:10.1177/1352458514556296

27. Karandikar NJ, Crawford MP, Yan X, Ratts RB, Brenchley JM, Ambrozak DR, Lovett- racke AE, Frohman EM, Stastny P, Douek DC, et al. Glatiramer acetate (Copaxone) therapy induces CD8 + T cell responses. J Clin Invest (2002) doi:10.1172/JCI200214380.Introduction

28. Bielekova B, Catalfamo M, Reichert-Scrivner S, Packer A, Cerna M, Waldmann TA, McFarland H, Henkart PA, Martin R. Regulatory CD56bright natural killer cells mediate immunomodulatory effects of IL-2Rα-targeted therapy (daclizumab) in multiple sclerosis. Proc Natl Acad Sci U S A (2006) doi:10.1073/pnas.0601335103

29. Stüve O, Marra CM, Jerome KR, Cook L, Cravens PD, Cepok S, Frohman EM, Phillips T, Arendt G, Hemmer B, et al. Immune surveillance in multiple sclerosis patients treated with natalizumab. Ann Neurol (2006) doi:10.1002/ana.20858

30. Saraste M, Irjala H, Airas L. Expansion of CD56Bright natural killer cells in the peripheral blood of multiple sclerosis patients treated with interferon-beta. Neurol Sci (2007) doi:10.1007/s10072-007-0803-3

31. Bielekova B, Richert N, Herman ML, Ohayon J, Waldmann TA, McFarland H, Martin R, Blevins G. Intrathecal effects of daclizumab treatment of multiple sclerosis. Neurology (2011) doi:10.1212/WNL.0b013e318239f7ef

32. Martínez-Rodríguez JE, López-Botet M, Munteis E, Rio J, Roquer J, Montalban X, Comabella M. Natural killer cell phenotype and clinical response to interferon-beta therapy in multiple sclerosis. Clin Immunol (2011) doi:10.1016/j.clim.2011.09.006

33. Börnsen L, Christensen JR, Ratzer R, Oturai AB, Sørensen PS, Søndergaard HB, Sellebjerg F. Effect of Natalizumab on Circulating CD4+ T-Cells in Multiple Sclerosis. PLoS One (2012) doi:10.1371/journal.pone.0047578

34. Welsh RM, Bahl K, Marshall HD, Urban SL. Type 1 interferons and antiviral CD8 T-Cell responses. PLoS Pathog (2012) doi:10.1371/journal.ppat.1002352

35. Matsui M, Mori KJ, Saida T, Akiguchi I, Kameyama M. The imbalance in CSF T cell subsets in active multiple sclerosis. Acta Neurol Scand (1988) doi: 10.1111/j.1600-0404.1988.tb05895.x

36. Hedlund G, Sandberg-Wollheim M, Sjögren HO. Increased proportion of CD4+ CDw29+ CD45R- UCHL-1+ lymphocytes in the cerebrospinal fluid of both multiple sclerosis patients and healthy individuals. Cell Immunol (1989) doi:10.1016/0008-8749(89)90388-2

37. Cepok S. Patterns of cerebrospinal fluid pathology correlate with disease progression in multiple sclerosis. Brain (2001) doi:10.1093/brain/124.11.2169

38. Oreja-Guevara C, Sindern E, Raulf-Heimsoth M, Malin JP. Analysis of lymphocyte subpopulations in cerebrospinal fluid and peripheral blood in patients with multiple sclerosis and inflammatory diseases of the nervous system. Acta Neurol Scand (2009) doi:10.1111/j.1600-0404.1998.tb01739.x

39. Mullen KM, Gocke AR, Allie R, Ntranos A, Grishkan I V., Pardo C, Calabresi PA. Expression of CCR7 and CD45RA in CD4 + and CD8 + subsets in cerebrospinal fluid of 134 patients with inflammatory and non-inflammatory neurological diseases. J Neuroimmunol (2012) doi:10.1016/jjneuroim.2012.04.017

40. Kowarik MC, Grummel V, Wemlinger S, Buck D, Weber MS, Berthele A, Hemmer B. Immune cell subtyping in the cerebrospinal fluid of patients with neurological diseases. J Neurol (2014) doi:10.1007/s00415-013-7145-2

41. Rodríguez-Martín E, Picón C, Costa-Frossard L, Alenda R, Sainz de la Maza S, Roldán E, Espiño M, Villar LM, Álvarez-Cermeño JC. Natural killer cell subsets in cerebrospinal fluid of patients with multiple sclerosis. Clin Exp Immunol (2015) doi:10.1111/cei.12580

42. Jing Y, Shaheen E, Drake RR, Chen N, Gravenstein S, Deng Y. Aging is associated with a numerical and functional decline in plasmacytoid dendritic cells, whereas myeloid dendritic cells are relatively unaltered in human peripheral blood. Hum Immunol (2009) doi:10.1016/j.humimm.2009.07.005

43. Krishnaraj R. Senescence and cytokines modulate the NK cell expression. Mech Ageing Dev (1997) doi:10.1016/S0047-6374(97)00045-6

44. Solana R, Alonso MC, Peña J. Natural killer cells in healthy aging. Exp Gerontol (1999) doi:10.1016/S0531-5565(99)00008-X

45. Mocchegiani E, Malavolta M. NK and NKT cell functions in immunosenescence. Aging Cell (2004) doi:10.1111/j.1474-9728.2004.00107.x

46. Zhang Y, Wallace DL, De Lara CM, Ghattas H, Asquith B, Worth A, Griffin GE, Taylor GP, Tough DF, Beverley PCL, et al. In vivo kinetics of human natural killer cells: The effects of ageing and acute and chronic viral infection. Immunology (2007) doi: 10.1111/j.1365-2567.2007.02573.x

47. Czesnikiewicz-Guzik M, Lee WW, Cui D, Hiruma Y, Lamar DL, Yang ZZ, Ouslander JG, Weyand CM, Goronzy JJ. T cell subset-specific susceptibility to aging. Clin Immunol (2008) doi:10.1016/j.clim.2007.12.002

48. Yani SL, Keller M, Melzer FL, Weinberger B, Pangrazzi L, Sopper S, Trieb K, Lobina M, Orrù V, Fiorillo E, et al. CD8+HLADR+ regulatory T cells change with aging: They increase in number, but lose checkpoint inhibitory molecules and suppressive function. Front Immunol (2018) doi:10.3389/fimmu.2018.01201

49. Li M, Yao D, Zeng X, Kasakovski D, Zhang Y, Chen S, Zha X, Li Y, Xu L. Age related human T cell subset evolution and senescence. Immun Ageing (2019) doi:10.1186/s12979-019-0165-8

50. Lee BW, Yap HK, Chew FT, Quah TC, Prabhakaran K, Chan GSH, Wong SC, Seah CC. Age- and Sex-Related changes in lymphocyte subpopulations of healthy Asian subjects: From birth to adulthood. Commun Clin Cytom (1996) doi:10.1002/(SICI)1097-0320(19960315)26:1<8::AID-CYT02>3.0.C0;2-E

51. Valiathan R, Ashman M, Asthana D. Effects of Ageing on the Immune System: Infants to Elderly. Scand J Immunol (2016) doi:10.1111/sji.12413

52. Abdullah M, Chai PS, Chong MY, Tohit ERM, Ramasamy R, Pei CP, Vidyadaran S. Gender effect on in vitro lymphocyte subset levels of healthy individuals. Cell Immunol (2012) doi:10.1016/j.cellimm.2011.10.009

53. Amadori A, Zamarchi R, De Silvestro G, Forza G, Cavatton G, Danieli GA, Clementi M, Chieco-Bianchi L. Genetic control of the CD4/CD8 T-cell ratio in humans. Nat Med (1995) doi:10.1038/nm1295-1279

54. Uppal SS, Verma S, Dhot PS. Normal values of CD4 and CD8 lymphocyte subsets in healthy indian adults and the effects of sex, age, ethnicity, and smoking. Cytometry (2003) doi:10.1002/cyto.b.10011

55. Kverneland AH, Streitz M, Geissler E, Hutchinson J, Vogt K, Boës D, Niemann N, Pedersen AE, Schlickeiser S, Sawitzki B. Age and gender leucocytes variances and references values generated using the standardized ONE-Study protocol. Cytom Part A (2016) doi:10.1002/cyto.a.22855

56. Garnett MR, Puget S, Grill J, Sainte-Rose C. Craniopharyngioma. Orphanet J Rare Dis (2007) doi: 10.1186/1750-1172-2-18

57. Cimino PJ, Zager M, McFerrin L, Wirsching HG, Bolouri H, Hentschel B, von Deimling A, Jones D, Reifenberger G, Weller M, et al. Multidimensional scaling of diffuse gliomas: application to the 2016 World Health Organization classification system with prognostically relevant molecular subtype discovery. Acta Neuropathol Commun (2017) doi:10.1186/s40478-017-0443-7

58. Sadighi Akha AA. Aging and the immune system: An overview. J Immunol Methods (2018) doi:10.1016/j.jim.2018.08.005

59. Esme M, Topeli A, Yavuz BB, Akova M. Infections in the Elderly Critically-Ill Patients. Front Med (2019) doi:10.3389/fmed.2019.00118

60. Mix E, Olsson T, Correale J, Kostulas V, Link H. CD4+, CD8+, and CD4- CD8- T Cells in CSF and Blood of Patients with Multiple Sclerosis and Tension Headache. Scand J Immunol (1990) 31:493-501. doi:10.1111/j.1365-3083.1990.tb02797.x

61. Barrau MA, Montalban X, Sáez-Torres I, Brieva L, Barberà N, Martínez-Cáceres EM. CD4+CD45RO+CD49d(high) cells are involved in the pathogenesis of relapsing- remitting multiple sclerosis. J Neuroimmunol (2000) doi:10.1016/S0165-5728(00)00357-X

62. Matsui M, Araya SI, Wang HY, Matsushima K, Saida T. Differences in systemic and central nervous system cellular immunity relevant to relapsing-remitting multiple sclerosis. J Neurol (2005) doi:10.1007/s00415-005-0778-z

63. Okuda Y, Okuda M, Apatoff BR, Posnett DN. The activation of memory CD4+ T cells and CD8+ T cells in patients with multiple sclerosis. J Neurol Sci (2005) doi:10.1016/j.jns.2005.02.013

64. Komori M, Blake A, Greenwood M, Lin YC, Kosa P, Ghazali D, Winokur P, Natrajan M, Wuest SC, Romm E, et al. Cerebrospinal fluid markers reveal intrathecal inflammation in progressive multiple sclerosis. Ann Neurol (2015) doi:10.1002/ana.24408

65. Magliozzi R, Howell O, Vora A, Serafini B, Nicholas R, Puopolo M, Reynolds R, Aloisi F. Meningeal B-cell follicles in secondary progressive multiple sclerosis associate with early onset of disease and severe cortical pathology. Brain (2007) doi:10.1093/brain/awm038

66. Howell OW, Reeves CA, Nicholas R, Carassiti D, Radotra B, Gentleman SM, Serafini B, Aloisi F, Roncaroli F, Magliozzi R, et al. Meningeal inflammation is widespread and linked to cortical pathology in multiple sclerosis. Brain (2011) doi:10.1093/brain/awr182

